# Risk stratification of childhood infection using host markers of immune and endothelial activation: a multi-country prospective cohort study in Asia (Spot Sepsis)

**DOI:** 10.1101/2025.02.03.25321543

**Authors:** Arjun Chandna, Constantinos Koshiaris, Raman Mahajan, Riris Adono Ahmad, Dinh Thi Van Anh, Suy Keang, Phung Nguyen The Nguyen, Sayaphet Rattanavong, Khalid Shams, Souphaphone Vannachone, Spot Sepsis Investigator Group, Mikhael Yosia, Naomi Waithira, Mohammad Yazid Abdad, Janjira Thaipadungpanit, Paul Turner, Phan Huu Phuc, Dinesh Mondal, Mayfong Mayxay, Bui Thanh Liem, Elizabeth A Ashley, Eggi Arguni, Rafael Perera-Salazar, Melissa Richard-Greenblatt, Yoel Lubell, Sakib Burza

## Abstract

**Background:** Circulating markers of immune and endothelial activation risk stratify infection syndromes agnostic to disease aetiology. However, their utility in children presenting from the community remains unclear.

**Methods:** This study recruited children aged 1-59 months presenting with community-acquired acute febrile illnesses to seven hospitals in Bangladesh, Cambodia, Indonesia, Laos, and Viet Nam. Clinical parameters and biomarker concentrations were measured at presentation. The outcome measure was death or receipt of vital organ support within two days of enrolment. Prognostic performance of endothelial (Ang-1, Ang-2, sFlt-1) and immune (CHI3L1, CRP, IP-10, IL-1ra, IL-6, IL-8, IL-10, PCT, sTNFR-1, sTREM-1, suPAR) activation markers, WHO Danger Signs, and two validated severity scores (LqSOFA, SIRS) was compared.

**Results:** 3,423 participants were recruited. 133 met the outcome (weighted prevalence: 0.34%; 95% CI 0.28-0.41). sTREM-1 exhibited highest prognostic accuracy (AUC 0.86; 95% CI 0.82-0.90), outperforming WHO Danger Signs (AUC 0.75; 95% CI 0.70-0.80; p < 0.001), LqSOFA (AUC 0.74; 95% CI 0.70-0.78; p < 0.001), and SIRS (AUC 0.63; 95% CI 0.58-0.68; p < 0.001). Discrimination of immune and endothelial activation markers was particularly strong for children who deteriorated later in the course of their illness. Compared to WHO Danger Signs, an sTREM-1-based triage strategy improved recognition of children at risk of progression to life-threatening infection (sensitivity: 0.80 vs. 0.72), while maintaining comparable specificity (0.81 vs. 0.79).

**Conclusions:** Measuring circulating markers of immune and endothelial activation may help earlier recognition of febrile children at risk of poor outcomes in resource-constrained community settings.

## INTRODUCTION

Whether to refer a febrile child to hospital is a challenging decision facing frontline community healthcare workers globally, particularly in resource limited and conflict affected settings.^1,2^ Each day, children who will develop severe disease are missed while referrals of illnesses suitable for community-based management incur avoidable cost for caregivers and health systems.^3,4^ In rural locations of many low- and middle-income countries, referral decisions are complex; influenced by poorly functioning health systems, limited referral infrastructure, and geographic, climatic, socioeconomic, and cultural factors.^5–7^

In under-resourced peripheral healthcare settings, the World Health Organization (WHO) recommends certain *Danger Signs* (convulsions, intractable vomiting, lethargy, or prostration), to identify febrile children requiring hospital referral.^8,9^ Yet, these suffer from considerable inter-observer variability and lack both sensitivity and specificity.^10,11^ Absence of data on children managed in the community setting renders their validity questionable.^12^ Better risk stratification tools for common childhood infections are needed.

Circulating markers of immune and endothelial activation have consistently demonstrated ability to risk stratify paediatric fever syndromes agnostic to disease aetiology.^13–15^ Elevated concentrations of these markers indicate loss of endothelial integrity and microvascular quiescence that contribute to disease progression, organ dysfunction, and death.^16–18^ They may be of particular value for identifying patients whose illness severity is not clinically apparent at presentation and who may be discharged and deteriorate at home.^19^ Whether these findings apply at the community setting in Asia is unknown: most research has included only hospitalised children, comprised single-site studies, and been conducted in locations where prevalent causes of infection and host susceptibility patterns differ.^13,20–23^

We report the first multi-country study of markers of immune and endothelial activation in children presenting from the community setting with acute febrile illnesses. Our objective was to determine whether presenting concentrations of these markers predict disease progression, thereby assessing their potential to identify children at risk of severe disease, relative to currently used clinical tools.

## METHODS

### Study design

Spot Sepsis was a multi-country, prospective, cohort study, which enrolled children (aged > 28 days and < 60 months) presenting with acute febrile illnesses to seven hospitals across Bangladesh, Cambodia, Indonesia, Lao PDR (Laos), and Viet Nam. Sites predominantly serving rural populations and providing a first point of contact with the formal healthcare sector were prioritised (appendix p2-3).

Patients presenting with a febrile illness (axillary temperature ≥ 37.5°C or < 35.5°C and/or history of fever in the preceding 24 hours) of ≤ 14 days duration were eligible for inclusion. Exclusion criteria were prior admission to any health facility during the current illness episode, receipt of > 15 minutes parenteral treatment before screening, presentation within 3 days of routine immunisations, trauma as the reason for attendance, and/or specific known comorbidities (chronic infection, immunosuppression, and/or active cardiorespiratory conditions). Participants could only be enrolled once.

Patients were screened at presentation to the outpatient and emergency departments during daytime working hours. Given high numbers of outpatients, consecutive enrolment of outpatients was not feasible and recruitment was stratified by admission status. Inpatients were enrolled consecutively. Outpatient recruitment was randomised using computer-generated random number tables, with the preceding week’s routinely collected hospital attendance data providing the sampling frame (Table S1; appendix p4-5)

Caregivers of all participants provided informed written consent. The study was prospectively registered on ClinicalTrials.gov (NCT04285021) and received ethical approval from the sponsors and ethical review boards in all participating countries (Table S2; appendix p6). MSF maintained a sponsor-investigator role for the study. The Wellcome Trust had no role in study design, data collection, data analysis, data interpretation, writing of the report, or decision to submit for publication.

### Data collection

Trained study personnel measured vital signs and anthropometrics, assessed clinical signs (including WHO Danger Signs), and collected venous blood samples and nasopharyngeal swabs at enrolment (Table S3; appendix p7). Demographics and perinatal, past medical, and illness histories were collected via interview with the participant’s caregiver and entered onto electronic case record forms using Android tablets via Open Data Kit Collect software. Variable selection was informed by systematic review of the literature.^24^ Prioritisation and standardisation followed guidance set out by the Pediatric Sepsis Predictors Standardization working group.^25^

Participants were followed-up on days 2 and 28 after enrolment, with additional follow-up on day 1 and at discharge for inpatients. Participants were provided with routine care by their treating clinician. When feasible, the study supported collection and processing of peripheral blood cultures at the discretion of the clinical team. Study monitoring was conducted by the Clinical Trials Support Group at the Mahidol-Oxford Tropical Medicine Research Unit (MORU) in Bangkok, Thailand.

### Selection of biomarkers and comparators

Biomarkers were selected following review of the literature and expert consultation (Table S4; appendix p8). Biomarkers useful for risk stratification in primary care, where the aetiology of infection is typically unknown at the time of assessment, must be predictive across a spectrum of pathogens. Hence, biomarkers with mechanistic links to final common pathways of severe febrile illness and sepsis were prioritised.^16,26–28^ Markers of endothelial activation included: angiopoietin-1 (Ang-1); angiopoietin-2 (Ang-2); and soluble fms-like tyrosine kinase-1 (sFlt-1; sVEGFR-1). Markers of immune activation included: chitinase-3-like protein-1 (CHI3L1); C-reactive protein (CRP); interferon-gamma-inducible protein-10 (IP-10; CXCL-10); interleukin-1 receptor antagonist (IL-1ra); interleukin-6 (IL-6); interleukin-8 (IL-8); interleukin-10 (IL-10); procalcitonin (PCT); soluble tumour necrosis factor receptor-1 (sTNFR-1); soluble triggering receptor expressed on myeloid cells-1 (sTREM-1); and soluble urokinase plasminogen activator receptor (suPAR).

Lactate, glucose, and haemoglobin were included as they are measurable using inexpensive rapid tests, familiar to many clinicians, have prognostic value,^24^ and promoted in paediatric sepsis guidelines.^29–31^

In addition to WHO Danger Signs, the Liverpool quick Sequential Organ Failure Assessment (LqSOFA) and Systemic Inflammatory Response Syndrome (SIRS) scores were selected as comparators (Table S5; appendix p9).^32,33^ LqSOFA is the most extensively studied age-adapted version of the widely-endorsed qSOFA sepsis screening tool for adults.^34^ It was developed specifically for triaging febrile children presenting from the community setting and outperformed other paediatric severity scores during external validation in Asia.^35^ Although the Phoenix Sepsis Score has superseded SIRS as the international consensus definition for paediatric sepsis, it does not yet offer a screening tool practicable in resource-limited frontline healthcare settings.^31^ Thus, SIRS was included as a widely-recognised paediatric sepsis screening tool.

### Laboratory procedures

Venous blood samples and nasopharyngeal swabs were processed immediately. Complete blood counts were performed on site. Peripheral blood cultures were processed at accredited in-country laboratories. Aliquots of whole blood, EDTA-plasma, fluoride-oxalate-plasma, and universal transport medium (UTM) were stored at −20°C or below. Samples were then transported at −80°C to the MORU laboratories in Bangkok, Thailand for further analysis and biobanking.

Biomarker concentrations were quantified in EDTA-plasma using the Simple Plex Ella microfluidic platform (ProteinSimple, San Jose, CA, USA) and suPARnostic ELISA (ViroGates, Denmark), as described in the appendix (Table S6; p10). Lactate (LACT2, Roche Diagnostics, Germany) and glucose (GLUC3, Roche Diagnostics, Germany) concentrations were quantified in fluoride-oxalate-plasma.

Nucleic acid was extracted from whole blood using the MagNA Pure 24 instrument and Total NA Isolation Kit (Roche Diagnostics, Indianapolis, IN, USA) according to manufacturer instructions. Whole blood viral (chikungunya, dengue, Japanese encephalitis, and Zika) and bacterial (*Leptospira* spp., *Orientia tsutsugamushi*, and *Rickettsia* spp.) targets were detected using laboratory developed real-time polymerase chain reaction (RT-PCR) multiplex assays.Respiratory pathogen targets were detected directly from nasopharyngeal swabs using the FilmArray RP2 panel (BioFire Diagnostics, Salt Lake City, UT, USA), with the exception of Cambodian samples, according to manufacturer protocols. Cambodian respiratory samples were processed for influenza A/B and respiratory syncytial virus (RSV) using the FTD FLU/HRSV assay (Siemens, Germany). All sites used an in-house developed multiplex RT-PCR assay for the detection of SARS-CoV-2 from nasopharyngeal swabs based on the E and N genes as described previously.^36^ Molecular targets were restricted to pathogens for which illness causality can be more confidently ascribed.^37^

### Outcomes

The outcome measure was development of severe febrile illness within two days of enrolment, defined as death and/or receipt of vital organ support (mechanical and/or non-invasive ventilation and/or inotropic therapy and/or renal replacement therapy).

Prespecified subgroup analyses included children with microbiologically-confirmed infections and different presenting clinical syndromes. Prognostic accuracy of the biomarkers and clinical assessment tools was explored across prediction horizons (< 4 hours, ≥ 4 hours, ≥ 24 hours, and ≥ 48 hours). These secondary analyses were planned to test the hypotheses that immune and endothelial activation markers would predict disease progression across different microbial aetiologies (i.e., were ‘pathogen agnostic’), and that value of biomarker measurements would be greatest in children whose illnesses progressed later after the point of presentation.^19,26,28^

### Sample size

Spot Sepsis had two main objectives prespecified in the study protocol: to examine the prognostic performance of individual host biomarkers and to develop a clinical prediction model.^38^ The methods of Riley et al. were followed to estimate the sample size required to build the clinical prediction model, reported separately, recognising that this would be adequate to evaluate the prognostic performance of individual host biomarkers.^39^

### Statistical analyses

Complete case analyses were used as missing data among the primary comparators (immune and endothelial activation markers and WHO Danger Signs) were few. Categorical and continuous variables were summarised using descriptive statistics and compared with the Wilcoxon rank sum test, Pearson’s X^2^ test, or Fisher’s exact test as appropriate. Site-specific outpatient weights were determined by estimating the proportion of all eligible outpatients recruited (Table S1; appendix p4-5). The prognostic accuracy of each biomarker, WHO Danger Signs, and clinical severity scores was quantified using the weighted area under the receiver operating characteristic curve (AUC). Probability weights were applied to adjust for unequal probabilities of selection in the sample, arising due to stratified recruitment. When evaluating combinations of characteristics, such as WHO Danger Signs and sTREM-1, a weighted logistic regression model was used to generate predicted probabilities, which were subsequently used to estimate the AUC.^40^

## RESULTS

### Study cohort

Between 5 March 2020 and 4 November 2022, 11,947 children were screened, of whom 3,995 were eligible (3,995/11,947; 33.4%) and 3,423 were recruited (572/3,995; 14.3% refusal rate). Eighteen participants were lost to follow-up (18/3,423; 0.5%) and excluded from further analyses (Figure S1; appendix p11).

Median age was 16.8 months (interquartile range [IQR] 8.7-31.0) and 60.0% of the cohort were male (2,029/3,405). Few participants had a known comorbidity (102/3,405; 3.0%). Approximately one in five children were wasted (weight-for-height z-score [WHZ] < −2; 585/3,393; 17.2%) and/or stunted (height-for-age z-score [HAZ] < −2; 664/3,401; 19.5%), and half of these were severely malnourished (WHZ and/or HAZ < −3). Median duration of illness prior to presentation was 3 days (IQR 2-4). The majority of children (2,333/3,405; 68.5%) lived within an hour of the hospital. 1,342 participants (1,342/3,405; 39.4%) had received care in the community at an earlier point in their illness: none had been admitted and 193 (193/3,405; 5.7%) had received parenteral treatment (Table S7; appendix p12). Table 1 shows presenting clinical data for the cohort. Additional information is provided in the appendix (Tables S8 and S9; p13-18).

**TABLE 1.** Presenting clinical characteristics, stratified by whether a child progressed to develop severe disease within two days of enrolment.

| Characteristic | Overall<br>N = 3,405<sup>1</sup> | Non-severe<br>N = 3,272<sup>1</sup> | Severe<br>N = 133<sup>1</sup> | p-value<sup>2</sup> |
| --- | --- | --- | --- | --- |
| **Demographics and background** |  |  |  |  |
| Age (months) | 16.8 (8.7, 31.0) | 17.3 (9.1, 31.3) | 4.9 (2.6, 17.3) | <0.001 |
| Male sex | 2,029 / 3,405 (60%) | 1,941 / 3,272 (59%) | 88 / 133 (66%) | 0.11 |
| Known comorbidity | 102 / 3,405 (3.0%) | 100 / 3,272 (3.1%) | 2 / 133 (1.5%) | 0.4 |
| Recent admission <sup>a</sup> | 429 / 3,394 (13%) | 412 / 3,262 (13%) | 17 / 132 (13%) | >0.9 |
| **Anthropometrics** |  |  |  |  |
| Weight-for-age z-score | -0.8 (-1.7, 0.0) | -0.8 (-1.7, 0.1) | -1.1 (-2.5, -0.2) | 0.004 |
| Wasted (WHZ < -2) <sup>b,*</sup> | 585 / 3,393 (17%) | 551 / 3,261 (17%) | 34 / 132 (26%) | 0.008 |
| Stunted (HAZ < -2) <sup>*</sup> | 664 / 3,401 (20%) | 629 / 3,268 (19%) | 35 / 133 (26%) | 0.044 |
| **Illness history** |  |  |  |  |
| Duration of illness (days) | 3.0 (2.0, 4.0) | 3.0 (2.0, 4.0) | 3.0 (2.0, 5.0) | 0.003 |
| Sought care prior to presentation | 1,753 / 3,405 (51%) | 1,681 / 3,272 (51%) | 72 / 133 (54%) | 0.5 |
| Travel time to study site ≤ 1 hour | 2,777 / 3,405 (82%) | 2,682 / 3,272 (82%) | 95 / 133 (71%) | 0.002 |
| **Presenting syndrome** |  |  |  |  |
| Upper respiratory tract infection | 1,121 / 3,405 (33%) | 1,082 / 3,272 (33%) | 39 / 133 (29%) | 0.4 |
| Lower respiratory tract infection | 1,347 / 3,405 (40%) | 1,261 / 3,272 (39%) | 86 / 133 (65%) | <0.001 |
| Diarrhoeal | 646 / 3,405 (19%) | 631 / 3,272 (19%) | 15 / 133 (11%) | 0.021 |
| Neurological | 430 / 3,405 (13%) | 416 / 3,272 (13%) | 14 / 133 (11%) | 0.5 |
| No focus | 527 / 3,405 (15%) | 519 / 3,272 (16%) | 8 / 133 (6.0%) | 0.002 |
| **Severity at presentation** |  |  |  |  |
| Any WHO Danger Sign present <sup>*</sup> | 1,607 / 3,398 (47%) | 1,512 / 3,266 (46%) | 95 / 132 (72%) | <0.001 |
| *Prostration* <sup>c</sup> | 240 / 3,405 (7.0%) | 192 / 3,272 (5.9%) | 48 / 133 (36%) | <0.001 |
| *Intractable vomiting* <sup>*</sup> | 687 / 3,401 (20%) | 645 / 3,268 (20%) | 42 / 133 (32%) | <0.001 |
| *Convulsions* <sup>*</sup> | 433 / 3,400 (13%) | 419 / 3,268 (13%) | 14 / 132 (11%) | 0.5 |
| *Lethargy* <sup>d,*</sup> | 825 / 3,399 (24%) | 764 / 3,267 (23%) | 61 / 132 (46%) | <0.001 |
| LqSOFA score <sup>*</sup> |  |  |  | <0.001 |
| 0 | 2,639 / 3,402 (78%) | 2,579 / 3,269 (79%) | 60 / 133 (45%) |  |
| 1 | 642 / 3,402 (19%) | 596 / 3,269 (18%) | 46 / 133 (35%) |  |
| 2 | 98 / 3,402 (2.9%) | 78 / 3,269 (2.4%) | 20 / 133 (15%) |  |
| 3 | 20 / 3,402 (0.6%) | 15 / 3,269 (0.5%) | 5 / 133 (3.8%) |  |
| 4 | 3 / 3,402 (<0.1%) | 1 / 3,269 (<0.1%) | 2 / 133 (1.5%) |  |
| Median LqSOFA score * | 0 (0, 0) | 0 (0, 0) | 1 (0, 1) | <0.001 |
| SIRS score * |  |  |  | 0.004 |
| 0 | 223 / 2,827 (7.9%) | 222 / 2,707 (8.2%) | 1 / 120 (0.8%) |  |
| 1 | 1,042 / 2,827 (37%) | 998 / 2,707 (37%) | 44 / 120 (37%) |  |
| 2 | 916 / 2,827 (32%) | 878 / 2,707 (32%) | 38 / 120 (32%) |  |
| 3 | 495 / 2,827 (18%) | 471 / 2,707 (17%) | 24 / 120 (20%) |  |
| 4 | 151 / 2,827 (5.3%) | 138 / 2,707 (5.1%) | 13 / 120 (11%) |  |
| Median SIRS score * | 2 (1, 2) | 2 (1, 2) | 2 (1, 3) | 0.006 |
| **Vital signs** |  |  |  |  |
| Heart rate * |  |  |  |  |
| 1 to 12 months (bpm) | 156.0 (140.0, 172.0) | 155.0 (140.0, 170.0) | 171.5 (157.5, 186.0) | <0.001 |
| 12 to 60 months (bpm) | 140.0 (127.0, 158.0) | 140.0 (126.0, 157.0) | 160.0 (140.0, 177.0) | <0.001 |
| Respiratory rate * |  |  |  |  |
| 1 to 12 months (bpm) | 45.0 (38.0, 55.0) | 44.0 (37.0, 53.0) | 59.5 (48.0, 66.0) | <0.001 |
| 12 to 60 months (bpm) | 36.0 (30.0, 42.0) | 36.0 (30.0, 42.0) | 39.0 (32.0, 55.0) | 0.006 |
| Oxygen saturation (%) * | 98.0 (97.0, 99.0) | 98.0 (97.0, 99.0) | 97.0 (95.0, 98.0) | <0.001 |
| Axillary temperature (°C) * | 37.6 (37.0, 38.3) | 37.6 (37.0, 38.3) | 37.6 (36.9, 38.3) | >0.9 |
| Prolonged capillary refill time <sup>e</sup> | 194 / 3,405 (5.7%) | 165 / 3,272 (5.0%) | 29 / 133 (22%) | <0.001 |
| Not alert <sup>f</sup> | 86 / 3,405 (2.5%) | 67 / 3,272 (2.0%) | 19 / 133 (14%) | <0.001 |

<sup>1</sup>Median (IQR); n / N (%); <sup>2</sup>Wilcoxon rank sum test; Pearson's Chi-squared test; Fisher's exact test
<sup>a</sup> overnight admission in last 6 months; <sup>b</sup> calculated in children 45-120cm; <sup>c</sup> unable to feed, sit, or stand when previously able; <sup>d</sup> abnormally sleepy and/or AVPU < A; <sup>e</sup> capillary refill time > 2 seconds; <sup>f</sup> AVPU < A.
\*Missing data: weight-for-height z-score (WHZ), n = 12; height-for-age z-score (LAZ), n = 4; WHO Danger Sign, n = 7; vomiting everything, n = 4; generalised seizures, n = 5; lethargy, n = 6; LqSOFA, n = 3; SIRS, n = 578; heart rate, n = 1; respiratory rate, n = 2; oxygen saturation, n = 205; axillary temperature, n = 1. Missingness for SIRS was greater, as complete blood counts were measured at the discretion of the treating clinical team.

### Outcomes

133 children met the outcome (133/3,405; 3.9%): 22 deaths and 111 survivors who required vital organ support (Bangladesh, n = 39; Cambodia, n = 36; Viet Nam 1, n = 32; Viet Nam 2, n = 26). The weighted outcome prevalence was 0.34% (95% confidence interval [CI] = 0.28-0.41; appendix p4-5). Young age, age-adjusted tachycardia, abnormal mental status, and bedside signs of poor peripheral perfusion and respiratory compromise were more common in participants who progressed to severe disease (Table 1; Table S8; appendix p13-14). Presence of a WHO Danger Sign at enrolment was associated with meeting the outcome, as were higher LqSOFA and SIRS scores (Table 1). Presenting plasma concentrations of the biomarkers stratified by outcome status are shown (Figure 1; Table S10; appendix p19).

**FIGURE 1.**
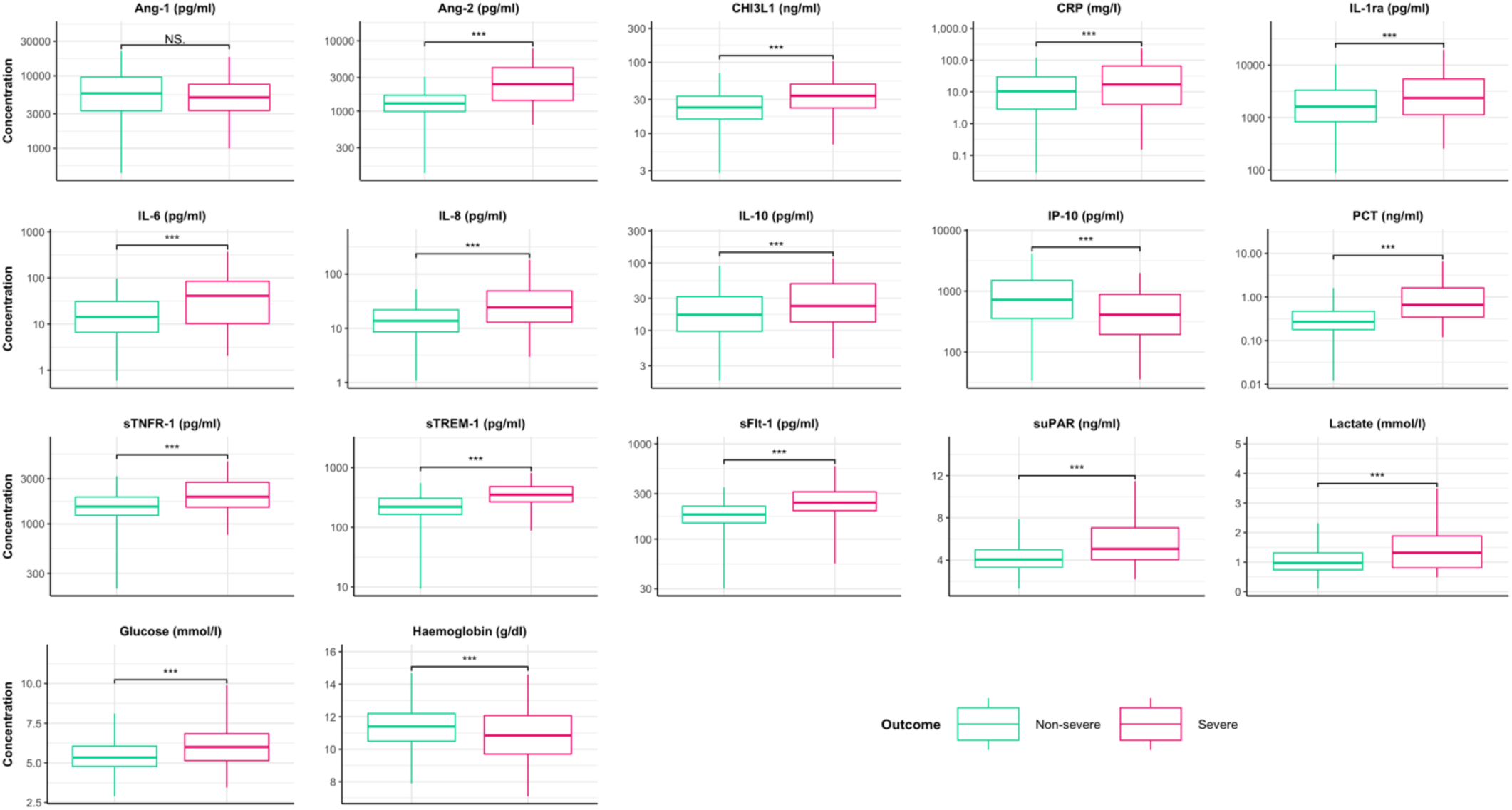
Presenting concentrations of circulating markers of endothelial and immune activation, stratified by whether a child progressed to develop severe disease within two days of enrolment. Box denotes middle 50% of the data, with the median indicated by the solid horizontal line. Upper and lower hinges denote 75^th^ and 25^th^ centile respectively. Whiskers extend from minimum to maximum value (hinge ± 1.5 times the interquartile range). Outliers not plotted to aid clarity. Presenting biomarker concentrations among children who progressed to severe disease (red) compared to children who did not progress to severe disease (green) using Wilcoxon rank sum test; NS = no statistically significant difference, *** = p < 0.001.

### Risk stratification

The predictive performance of each circulating marker is presented in Figure 2a, alongside the performance of WHO Danger Signs and the clinical severity scores. sTREM-1 showed best prognostic accuracy (AUC 0.86; 95% CI 0.82-0.90), demonstrating superior ability to discriminate children who would progress to severe disease, compared to other circulating markers, WHO Danger Signs (AUC 0.75; 95% CI 0.70-0.80; p < 0.001), and the clinical severity scores (LqSOFA: AUC 0.74; 95% CI 0.70-0.78; p < 0.001; SIRS: AUC 0.63; 95% CI 0.58-0.68; p < 0.001). Combining WHO Danger Signs with sTREM-1 (AUC 0.88; 95% CI 0.85-0.91) did not improve performance over sTREM-1 alone (p = 0.24; Figure 2b)

**FIGURE 2.**
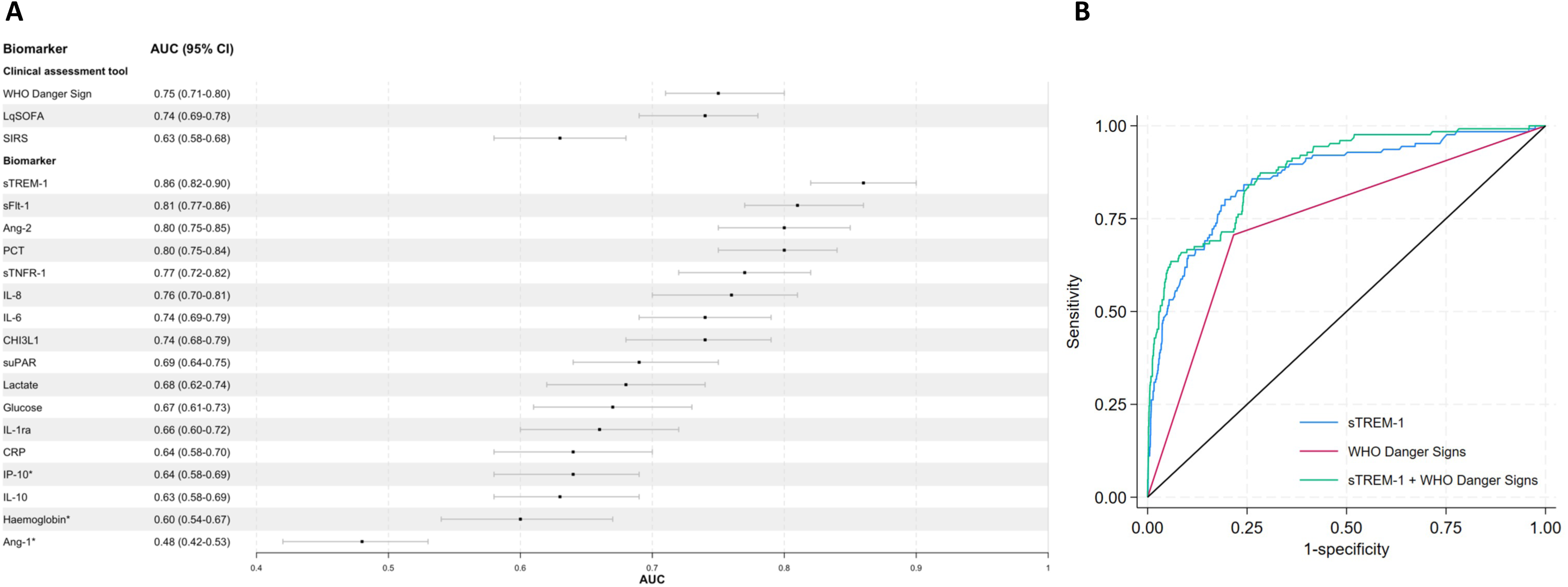
A: Prognostic performance of clinical assessment tools and circulating markers of endothelial and immune activation to predict progression to severe disease within two days of enrolment. B: Prognostic performance of WHO Danger Signs and sTREM-1, alone and in combination, to predict progression to severe disease within two days of enrolment. A: Solid square = point estimate for AUC; error bars = 95% CI. *Reciprocal concentration used, as lower biomarker concentrations known to be indicative of more severe disease. B: Receiver operating characteristic curves for WHO Danger Signs (red) AUC = 0.75 (95% CI 0.70-0.80); sTREM-1 (blue) AUC = 0.86 (95% CI 0.82-0.90); and sTREM-1 plus WHO Danger Signs (green) AUC = 0.88 (95% CI 0.85-0.91).

Sensitivity and specificity of WHO Danger Signs for recognising children who would progress to severe disease was 0.72 (95% CI 0.66-0.79) and 0.79 (95% CI 0.76-0.82), respectively. sTREM-1 concentrations selected to provide equivalent sensitivity or specificity, improved classification (Table 2). Using the Youden index to identify a sTREM-1 threshold for triage resulted in a sensitivity of 0.80 (95% CI 0.73-0.85) and specificity of 0.81 (95% CI 0.78-0.83). At the current outcome prevalence (0.34%), compared to using WHO Danger Signs, sTREM-1-based triage would identify one additional child who would progress to life-threatening infection for every ∼3,000 children tested, without compromising specificity (increasing false positives).

**TABLE 2.** Sensitivity and specificity of WHO Danger Signs and sTREM-1 for recognition of children who progressed to severe disease within two days of enrolment.

| Parameter | Threshold | Method for selection | Sensitivity (95% CI) | Specificity (95% CI) |
| --- | --- | --- | --- | --- |
| **WHO Danger Signs** | Presence/absence | NA | 0.72 (0.66-0.79) | 0.79 (0.76-0.82) |
| **sTREM-1** | 279 pg/ml | Fixed at sensitivity of WHO Danger Signs | 0.72 (0.65-0.79) | 0.83 (0.81-0.86) |
| **sTREM-1** | 257 pg/ml | Fixed at specificity of WHO Danger Signs | 0.80 (0.74-0.86) | 0.79 (0.76-0.82) |
| **sTREM-1** | 261 pg/ml | Youden index | 0.80 (0.73-0.85) | 0.81 (0.78-0.83) |

Recognising that the relative importance of sensitivity and specificity is context dependent, the performance of sTREM-1 was compared to WHO Danger Signs by evaluating specificity at the sTREM-1 concentration that equated to the sensitivity of WHO Danger Signs, and then by evaluating sensitivity at the sTREM-1 concentration that equated to the specificity of WHO Danger Signs. Finally, the Youden index was used to identify the sTREM-1 concentration which maximised accuracy (the optimal trade-off between sensitivity and specificity, assuming both were of equal importance).

### Prognostication in microbiologically-confirmed infections

A microbiological cause for infection was confirmed in 898 children (898/3,405; 26.4%): 429 RSV, 164 arboviral infections (109 dengue, 47 chikungunya, 8 Zika); 146 influenza (87 influenza A, 59 influenza B); 81 SARS-CoV-2; 59 human metapneumovirus; 19 bacteraemias (Table S11; appendix p20); 9 rickettsial infections (6 *Rickettsia* spp., 3 *Orientia tsutsugamushi*); 9 pertussis (8 *Bordetella parapertussis*, 1 *Bordetella pertussis*); 4 leptospirosis; 3 *Chlamydia pneumoniae;* and 3 *Mycoplasma pneumoniae*. Thirty four children had co-infections with two pathogens. Full details are provided in the appendix (Table S11; p20). Amongst participants with microbiologically-confirmed infections, prognostic accuracy of the circulating markers, WHO Danger Signs, and LqSOFA was largely unchanged (Table S12; appendix p21), with sTREM-1 providing best discrimination (AUC 0.88; 95% CI 0.83-0.94). Few participants had confirmed bacterial infections (n = 47), precluding comparison of prognostic performance between children with viral and bacterial infections.

### Syndrome-specific prognostication

In children whose presentations met WHO-pneumonia criteria (cough and/or difficult breathing with age-adjusted tachypnoea and/or chest indrawing),^8^ prognostic accuracy of sTREM-1 (AUC 0.84; 95% CI 0.78-0.94; Table S12; appendix p21) was matched by two markers of endothelial activation, Ang-2 (AUC 0.85; 95% CI 0.79-0.91) and sFlt-1 (AUC 0.84; 95% CI 0.77-0.90). Discrimination of the clinical assessment tools was poorer: WHO Danger Signs (AUC 0.58; 95% CI 0.50-0.65; p < 0.001); LqSOFA (AUC 0.72; 95% CI 0.66-0.78; p < 0.001); and SIRS (AUC 0.62; 95% CI 0.53-0.70; p < 0.001). The remaining outcome events were dispersed across clinical syndromes, precluding additional syndrome-specific analyses. Aggregate results for all non-respiratory presentations are included in the appendix (Table S12; p21).

### Prognostication across prediction horizons

Extending the prediction horizon to include all cases of severe febrile illness occurring during follow-up (up to day 28), identified an additional 10 children (2 deaths and 8 survivors who required vital organ support). Data for time-stratified analyses were available for 139/143 (97.2%) children: 56 met the outcome within 4 hours, 83 after more than 4 hours, 42 after more than 24 hours, and 21 after more than 48 hours from enrolment. For most circulating markers there was a trend to improved discrimination at distal prediction horizons (Figure 3). WHO Danger Signs performed consistently across prediction horizons. Performance of the clinical severity scores was better for children whose illnesses progressed soon after enrolment. sTREM-1 remained superior to other markers and clinical assessment tools across all horizons, demonstrating an AUC of 0.94 (95% CI 0.89-0.98) for discrimination of children who progressed to severe disease more than 48 hours after presentation.

**FIGURE 3.**
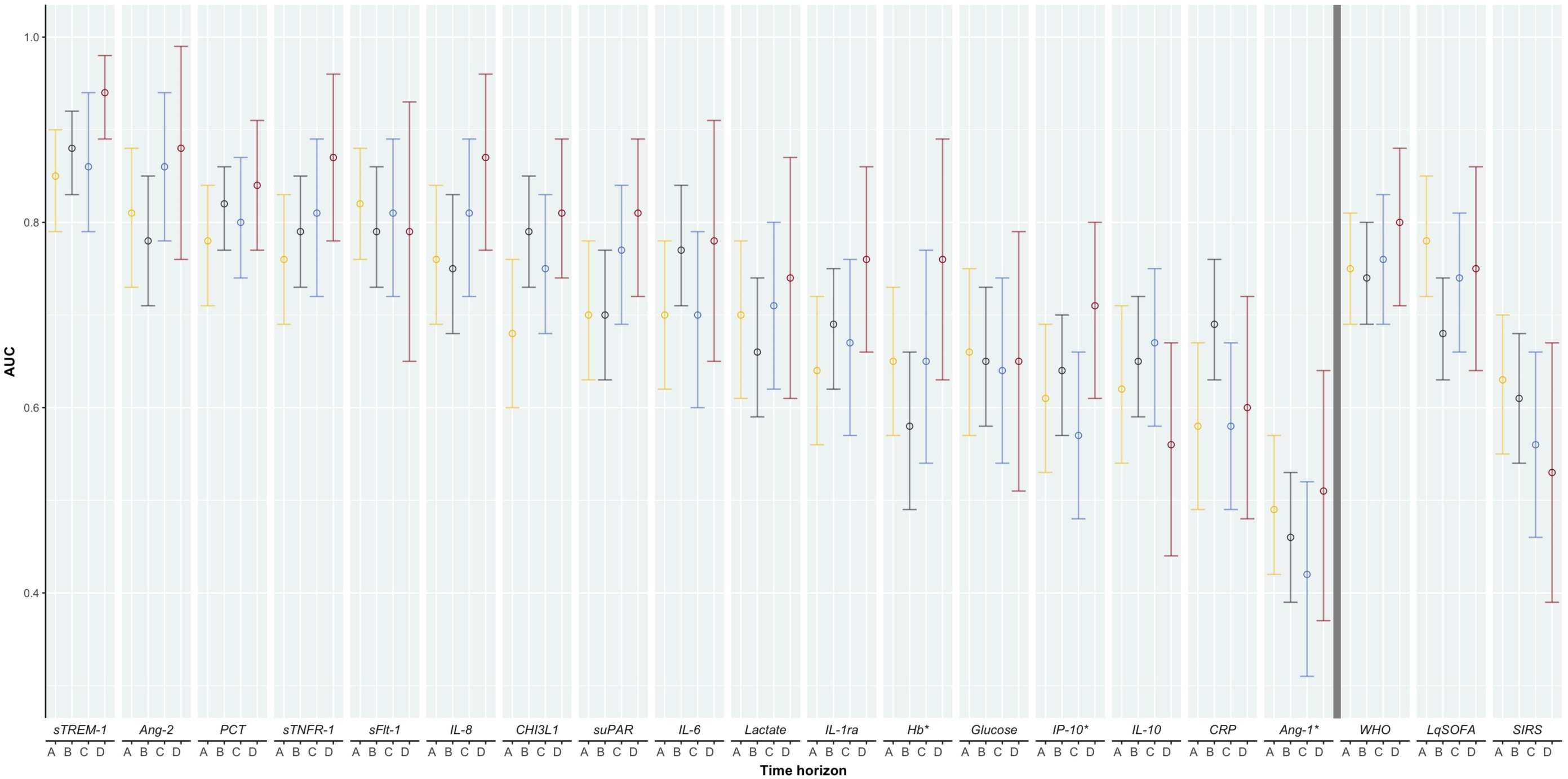
Prognostic performance of clinical assessment tools and circulating markers of endothelial and immune activation to predict progression to severe disease across different time horizons. Prediction horizons (events/non-events): A (yellow) = < 4 hours (56/3,319); B (grey) = ≥ 4 hours (83/3,236); C (blue) = ≥ 24 hours (42/3,236); D (red) = ≥ 48 hours (21/3,236). Participants who met the outcome by horizon A are excluded from horizon B, C, and D analyses. Participants who met the outcome prior to 24 or 48 hours are excluded from horizon C and D analyses respectively. Biomarkers presented to the left of solid vertical line, clinical assessment tools presented to the right, both in descending order of mean AUC across prediction horizons. Open circle = point estimate for AUC; error bars = 95% CI.*Reciprocal concentration used, as lower biomarker concentrations known to be indicative of more severe disease.

### Sensitivity analyses

sTREM-1 maintained prognostic accuracy and outperformed the clinical assessment tools across sites (AUCs 0.84-0.89; Table S13; appendix p22-23). Sensitivity analyses excluding the northern Viet Nam site (n = 612), which departed from the ideal rural target site profile and where outpatient weighting was derived using different methodology (Tables S1 and S13; appendix p4-5 and p22-23), did not affect the results. Finally, in a sensitivity analysis restricted to children who had not received parenteral treatment at the study site prior to baseline data or sample collection (3,037/3,045; 89.2%), sTREM-1 remained the best prognostic indicator (AUC 0.82; 95% CI 0.76-0.88; Tables S14a and S14b; appendix p24-25).

## DISCUSSION

In this large and geographically diverse Asian study investigating circulating markers of immune and endothelial activation for the risk stratification of unselected febrile children presenting from community settings, we found sTREM-1 to be consistently superior to WHO Danger Signs (the current standard of care) and LqSOFA (a validated clinical severity score) across prediction horizons, sites, presenting clinical syndromes, and in participants with microbiologically-confirmed infections.

Previous studies highlight the promising prognostic performance of sTREM-1. ^13,20,21,23,41,42^ However, these focussed exclusively on hospitalised children. Our results provide the first definitive evaluation at the community level, where need for better triage tools is most urgent.^1,43^ We identified sites serving as the first point of presentation for rural populations, enrolled unselected febrile children including outpatients, recruited participants immediately upon presentation, excluded children admitted elsewhere prior to screening, and adopted an analysis strategy ensuring an outcome prevalence reflective of community care settings.^44,45^

Our findings support the evidence that certain circulating markers of endothelial and immune activation are pathogen agnostic, reflecting final common pathways to severe infection.^15,26,28^ Thus, they are attractive candidates for risk stratification in primary care, where the cause of infection is typically unknown at the time of triage and recognising which child’s illness is likely to progress remains a major clinical challenge. Endothelial dysfunction has been demonstrated in ambulatory children with infection.^46^ However, until now it was unclear whether concentrations of these markers would be elevated sufficiently early in the natural history of infection to permit their use for risk stratification at the community level. The results of this study, in conjunction with two previously published smaller community-based studies, suggest that this approach warrants further attention.^19,27^

The results of our study are consistent with a study of 507 febrile adults conducted in Tanzanian outpatient clinics, which reported an AUC for sTREM-1 of 0.87 (95% CI 0.81-0.92) for predicting death within 28 days.^27^ In a study of children with pneumonia presenting to a primary care clinic on the Thailand-Myanmar border, Ang-2 demonstrated best prognostic performance (AUC 0.81; 95% CI 0.74-0.87) to predict supplemental oxygen requirement, whereas sTREM-1 did not show discriminatory value (AUC 0.56; 95% CI 0.49-0.63).^19^ In part, this may relate to the focus on pneumonia: Ang-2 was the top-performing marker amongst children with pneumonia in our study, although performance of sTREM-1 also remained comparable. Alternatively, the contrasting findings may be explained by the more proximal endpoint (supplemental oxygen requirement vs. vital organ support and death) or pre-analytical differences in sample matrix or storage conditions, which are known to influence biomarker concentrations.^47^

This is the first multi-country study investigating circulating markers of immune and endothelial activation in childhood infection. Other key strengths include: a study design which maximised relevance for community settings as detailed above; the inclusion of a prespecified panel of biomarkers compiled based on existing literature and underpinned by mechanistic links to sepsis pathophysiology, which lends face validity to the findings; simultaneous quantification of multiple markers in a central laboratory to ensure comparability of findings; and recruitment across seven sites in five countries for over 30 months, which improves geographic and seasonal generalisability.

There are several limitations. Despite steps taken to optimise external validity to community settings, inherent differences between patients presenting to rural hospital outpatient departments and primary care facilities will remain. Amongst children progressing to severe febrile illness, median time to developing severe disease was 6 hours (IQR 2-30), indicating a level of severity at presentation that may not be replicated in some community care settings. Nevertheless, time-stratified analyses demonstrate that discrimination of most circulating markers was strongest in children who progressed to severe disease later; a finding not observed for the comparator clinical assessment tools. This suggests that biomarkers may be of greatest value in patients whose illness severity is not clinically apparent at presentation and who are at risk of being sent home and deteriorating. This is consistent with previous work in childhood pneumonia and has implications for operationalising biomarker tests.^19^ Our analyses were structured such that every child would receive a biomarker test. Whilst this may be appropriate in certain settings,^1,48^ in others a point-of-care test would likely be used selectively on children for whom decisions to refer are borderline. Future work must explore different strategies for integrating biomarker testing into patient triage and compare the cost-effectiveness of different approaches.

Our outcome measure was selected as it is unlikely to be influenced by factors other than disease severity. Small amounts of outcome misclassification can substantially impact estimates of predictor performance.^49^ Nevertheless, predictors of severe disease may not generalise to more proximal outcomes. Studies in Covid-19 and childhood pneumonia indicate that sTREM-1 concentrations do not predict supplemental oxygen requirement as accurately as they do mortality.^19–21,50,51^ This underscores the importance of including a range of outcomes when evaluating the performance of predictors in primary care.

The impact of the Covid-19 pandemic must be considered. The majority of children (2,861/3,405; 84.0%) were tested for SARS-CoV-2 and few (81/2,861; 2.8%) found to be infected. Findings should not be biased towards biomarkers implicated in this specific infection. Nevertheless, health systems and care-seeking pathways were substantially impacted during the pandemic, with both attendance rates and the proportion of patients with severe outcomes generally lower than anticipated based on pre-pandemic baseline data. In particular, no severe outcomes were observed at the Indonesia or Laos sites. It will be important to assess the generalisability of our results in non-pandemic times.

Concentrations of circulating markers of immune and endothelial activation predict disease severity across a spectrum of common childhood infections. We demonstrate that these findings may be applicable at the community level, where need for better risk stratification tools is most urgent. Amongst the markers studied, sTREM-1 holds most potential, demonstrating improved sensitivity and specificity compared to the existing standard of care. Future work should focus on validating these findings, explore different approaches for integrating biomarker testing into patient triage, and assess cost-effectiveness. Priority should be given to biomarkers that are harbingers for disease progression and facilitate earlier recognition of patients in whom illness severity is not yet clinically apparent at presentation. Ultimately, point-of-care tests for the most promising biomarkers must be developed if the clinical utility of biomarker-based triage strategies is to be assessed in definitive randomised controlled trials.

## CONTRIBUTORS

AC, CK, RAA, PT, MM, EAA, EA, RPS, MRG, YL, and SB conceptualised the study. AC, DTVA, SK, PNTN, SR, KS, SV, PHP, DM, BTL, and EA acquired the data. AC, RM, and MRG curated the data. AC, CK, RM, and RPS did the formal analysis. YL and SB acquired funding. AC wrote the original draft of the manuscript. AC, CK, RM, RAA, DTVA, SK, PNTN, SR, KS, SV, PT, PHP, DM, MM, BTL, EAA, EA, RPS, MRG, YL, and SB reviewed and edited the manuscript. AC, CK, RM, MRG, YL, and SB verified the underlying data.

## DATA SHARING

De-identified, individual participant data from this study will be available to researchers whose proposed purpose of use is approved by the data access committees at *Médecins Sans Frontières* and the Mahidol-Oxford Tropical Medicine Research Unit. Enquiries or requests for the data may be sent to. Researchers interested in accessing biobanked samples should contact the corresponding author who will coordinate with the Spot Sepsis Sample Use Committee.

## ACKNOWLEDGEMENTS

We are extremely grateful to all the participants and their caregivers, and the laboratory and research staff at the participating sites and MORU laboratories in Bangkok for specimen processing, data management, and study monitoring. We acknowledge the support of the Mérieux Foundation who provided laboratory consumables and equipment for some of the respiratory specimen assays. We also acknowledge the valuable support and guidance of Nicholas J White (MORU) at key points during the study.

