## Appendix for "Risk stratification of childhood infection using host markers of immune and endothelial activation: a multi-country prospective cohort study in Asia (Spot Sepsis)"

### SUPPLEMENTARY APPENDIX

### 1. Details of the study sites.

Sites located outside major urban cities were proactively approached. As part of the shortlisting process, sites were asked to indicate the proportion of their patient population residing in a rural location and the proportion of children using the hospital as a first point of contact with the formal healthcare sector. Where possible, routinely collected data were used to inform site selection. If these were not available, local clinicians and hospital administrators were asked to provide their best estimate.

Due to the disruptions caused by the Covid-19 pandemic, site activation was delayed and recruitment proceeded slower than anticipated at all sites. After 18 months of recruitment, a decision was taken to identify an additional site in order to boost recruitment, ensure sufficient outcome events, and safeguard viability of the study. The second Viet Nam site was subsequently identified and brought online in December 2021, acknowledging that it departed from the desired rural site target profile established at the outset of the study. A sensitivity analysis excluding this site was prespecified in order to address this.

**Bangladesh**
*Recruitment period: 18/03/2021 – 27/04/2022*
Goyalmara Mother and Child Hospital is a non-governmental hospital managed by Médecins Sans Frontières located in Cox's Bazaar, Chattogram Division, in eastern Bangladesh, which predominantly provides health services to the forcibly displaced Rohingya refugee population. The hospital provides primary and secondary care, with approximately 20,000 outpatient attendances and 4,000 inpatient admissions annually. The 12-bed high-dependency unit provides non-invasive ventilation and inotropic therapy. There is a basic on-site diagnostic laboratory.

**Cambodia**
*Recruitment period: 05/03/2020 – 24/02/2022*
Angkor Hospital for Children is a non-governmental paediatric hospital located in Siem Reap province, northern Cambodia. The hospital provides primary-to-tertiary care, with approximately 80,000 outpatient attendances and 3,000 inpatient admissions annually. The 14-bed intensive care unit provides non-invasive ventilation, mechanical ventilation, inotropic therapy, and peritoneal dialysis. There is an on-site diagnostic microbiology laboratory (ISO15189 accredited since 23 November 2023).

**Indonesia**
*Recruitment period: 22/03/2021 – 22/04/2022*
Rumah Sakit Umum Daerah Wates is a government district hospital located in Yogyakarta province, Indonesia. The hospital provides primary and secondary care, with approximately 40,000 outpatient attendances and 4,000 inpatient admissions annually. The 8-bed intensive care unit provides non-invasive ventilation, mechanical ventilation, inotropic therapy, and renal replacement therapy. Basic microscopy is available on-site. Culture-based microbiology is available via nearby private laboratories.

**Laos 1**
*Recruitment period: 10/09/2020 – 30/08/2021*
Salavan Provincial Hospital is the government provincial hospital for Salavan, a predominantly rural province in southern Laos. The hospital provides primary-to-tertiary care, with approximately 65,000 outpatient attendances and 12,000 inpatient admissions annually. The 3-bed paediatric intensive care unit provides non-invasive ventilation, mechanical ventilation, and inotropic therapy. There is an on-site diagnostic microbiology laboratory.

**Laos 2**
*Recruitment period: 21/01/2021 – 26/08/2021*

Savannakhet Provincial Hospital is the government provincial hospital for Savannakhet, a predominantly rural province in southern Laos. The hospital provides primary-to-tertiary care, with approximately 7,000 outpatient attendances and 3,000 inpatient admissions annually. The 22-bed intensive care unit provides non-invasive ventilation, mechanical ventilation, and inotropic therapy. There is an on-site diagnostic microbiology laboratory.

**Viet Nam 1**
*Recruitment period: 10/05/2021 – 28/10/2022*
Dong Nai Children's Hospital is the government provincial paediatric hospital for Dong Nai province in southern Viet Nam. The hospital provides primary-to-tertiary care, with approximately 80,000 outpatient attendances and 7,000 inpatient admissions annually. The 30-bed intensive care unit provides non-invasive ventilation, mechanical ventilation, inotropic therapy, and renal replacement therapy. There is an on-site diagnostic microbiology laboratory.

**Viet Nam 2**
*Recruitment period: 08/12/2021 – 04/11/2022*
Viet Nam National Children's Hospital is the government national paediatric hospital, located in Ha Noi. The hospital provides primary-to-quaternary care, with approximately 1,200,000 outpatient attendances and 110,000 inpatient admissions annually. The 40-bed intensive care unit provides non-invasive ventilation, mechanical ventilation, inotropic therapy, and renal replacement therapy. There is an on-site diagnostic microbiology laboratory.

### 2. Table S1: Derivation of site-specific outpatient weights.

| | Bangladesh | Cambodia | Laos 1 | Laos 2 | Viet Nam 1 | Viet Nam 2 | Total |
|---|---|---|---|---|---|---|---|
| **Screening week data** | | | | | | | |
| Number of screening weeks | 3 | 4 | 2 | 1 | 2 | - | - |
| Patients screened (A) | 1,556 | 1,872 | 204 | 39 | 899 | - | - |
| Patients eligible (B) | 320 | 346 | 71 | 4 | 180 | - | - |
| Proportion eligible (B/A = C) | 0.21 | 0.19 | 0.35 | 0.10 | 0.20 | 0.20 | - |
| **Routine hospital data** | | | | | | | |
| Outpatient attendance during study (D) | 28,613 | 59,853 | 5,403 | 1,278 | 48,296 | 36,854 | - |
| Estimated number eligible (C*D = E) | 5,867 | 11,073 | 1,891 | 128 | 9,660 | 7,371 | - |
| **Study data** | | | | | | | |
| Number of outpatients recruited (F) | 175 | 183 | 87 | 29 | 147 | 147 | - |
| Outpatient weighting (1 : E/F) | 1 : 34 | 1 : 64 | 1 : 22 | 1 : 5 | 1 : 66 | 1 : 50 | - |
| **Outcome prevalence** | | | | | | | |
| Unweighted (95% CI) | 7.1% (5.2-9.5) | 4.4% (3.2-5.9) | - | - | 3.4% (2.4-4.7) | 3.5% (2.4-5.0) | **3.9% (3.3-4.6)** |
| Weighted (95% CI) | 0.64% (0.45-0.90) | 0.30% (0.20-0.42) | - | - | 0.31% (0.21-0.45) | 0.39% (0.26-0.59) | **0.34% (0.28-0.41)** |

Due to high numbers of outpatients, consecutive enrolment of outpatients was not feasible and recruitment was stratified by admission status, with consecutive inpatient enrolment and randomised outpatient recruitment. In order to account for this stratification and ensure relevance of the results to community settings where prevalence of severe disease is lower,[1,2] the outpatient strata had to be weighted in the analyses. For one week every 4-6 months, outpatient recruitment was paused and consecutive attendances screened for eligibility. These screening week data were triangulated with routinely collected hospital attendance data to estimate the total number of eligible outpatients presenting to each site during the recruitment period, in order to determine the weights to be used in the analyses.

Accordingly, screening weeks were planned at each site where outpatient recruitment was randomised. At study inception outpatient recruitment was randomised at all sites. Following the start of the Covid-19 pandemic, attendance rates at the site in Indonesia decreased substantially, such that consecutive outpatient recruitment became possible. The switch to consecutive recruitment in Indonesia occurred on 10/04/2021 (three weeks after site initiation), covering 90.4% (66/73) of outpatient recruitment at that site. As consecutive recruitment was used for the majority of outpatient recruitment in Indonesia, no weighting was necessary. However, to align with the methodology used at the other sites, where the estimated number of eligible outpatients presenting to the study site during the recruitment period (E) assumed no refusals, the Indonesian outpatient data were weighted by the observed outpatient refusal rate (62/135; 46%), to provide a weighting of 1:2 for the Indonesian outpatient data.

The second Viet Nam site joined the study on 8 December 2021 in order to boost recruitment, which had been delayed by the Covid-19 pandemic (appendix p2-3). Due to limited remaining resources at this stage of the study, it was not possible to recruit outpatients at this site. Therefore, outpatient data from the first Viet Nam site are used as a proxy. Weighting of these data for the second Viet Nam site was performed using the ratio of inpatients recruited at each Vietnamese site (612:802) and the total number of outpatient attendances at the first Viet Nam site (48,296), to estimate the number of outpatients that would have presented to the second Viet Nam site: 48,296 * (612/802) = 36,854. Assuming the same proportion (0.20) of eligible outpatients at both Viet Nam sites, the number of eligible outpatients presenting to the second Viet Nam site was estimated as: 36,854 * 0.20 = 7,371. This number was then used to determine the weighting to apply to the outpatient data from the first Viet Nam site, as a proxy for outpatient data at the second Viet Nam site: 7,371/147 = 50. The key assumptions underlying this are that the ratio of inpatients to outpatients, proportion of eligible outpatients, and profile of outpatients are similar at both Vietnamese sites. A sensitivity analysis excluding data from the second Viet Nam site produced similar results to the main analysis.

### 3. Table S2: Ethical approvals.

| Ethical Review Board | Approval Reference | Country |
|---|---|---|
| Médecins Sans Frontières Ethical Review Board | MSF ERB 1967 | NA |
| Oxford Tropical Medicine Research Committee | OxTREC 59-19 | NA |
| International Centre for Diarrhoeal Disease Research | PR-200006 | Bangladesh |
| Angkor Hospital for Children Research Committee | 01296/19AHC | Cambodia |
| National Ethics Committee for Health Research | 264/NECHR | Cambodia |
| Medical and Health Research Ethics Committee | KE/FK/1397/EC/2019 | Indonesia |
| National Ethics Committee for Health Research | 051/NECHR | Laos |
| University of Medicine and Pharmacy at Ho Chi Minh City | 818/HDDD-DHYD | Viet Nam |
| Ethics Committee for Biomedical Research | VNCH-RICH-2021-77 | Viet Nam |

### 4. Table S3: Methodology for collection of predictor variables.

| Variable | Methodology |
|---|---|
| Respiratory rate | Manual count for 60 seconds using clicker counter and timer |
| Heart rate | Massimo Rad-5v pulse oximeter with paediatric and neonatal probes |
| Oxygen saturation | Massimo Rad-5v pulse oximeter with paediatric and neonatal probes |
| Axillary temperature | Digital thermometer: operating range 32.0-42.9°C; accuracy ± 0.1°C |
| Capillary refill time | Pressure applied to sternum for 5 seconds |
| Length / Height | Médecins Sans Frontières height and length board |
| Weight | Seca 877 scale with mother-and-child function; accuracy ± 50g |
| Mid-upper arm circumference | Médecins Sans Frontières traffic light MUAC tape |
| Mental status | Alert Voice Pain Unresponsive (AVPU) scale |
| WHO Danger Signs | WHO IMCI Distance Learning course: https://iris.who.int/handle/10665/104772 |

5. **Table S4: Existing evidence supporting selection of endothelial and immune activation markers.**

| Biomarker | Overview of supportive data |
|---|---|
| **Endothelial activation** | |
| **Ang-1 and -2** | Supportive data from Asia/SSA/Europe in children/adults, that increases in Ang-2, decreases in Ang-1, and/or the Ang-2:1 ratio predicts mortality in pneumonia, malaria, SBI, and all-cause febrile illnesses,[3-12] and supplemental oxygen requirement in children with pneumonia in Asia.[13] |
| **sFlt-1** | Supportive data from SSA that increases in sFlt-1 predict mortality in children hospitalised with pneumonia, severe malaria, and all-cause febrile illnesses, and adults with all-cause febrile illnesses.[3,7,8,11,14] |
| **Immune activation** | |
| **CHI3L1** | Supportive data from SSA that increases in CHI3L1 predict mortality in children hospitalised with pneumonia and all-cause febrile illnesses, and adults with all-cause febrile illnesses.[3,7,8] |
| **CRP** | Although there is limited supportive evidence for the use of CRP as a prognostic marker for disease severity,[15] as it is the most widely studied biomarker in the region, and numerous point-of-care tests already exist, further evaluation is warranted. |
| **IL-1ra** | Supportive data that increases in IL-1ra are associated with severity in children with meningococcal disease, adults with SARS-CoV-2 infection, and predict need for longer antibiotic duration in children with febrile lower respiratory tract infections.[16-18] |
| **IL-6** | Supportive data from India that increases in IL-6 are predictive of mortality in children with dengue;[19] in Switzerland, supportive data that increases in IL-6 predict need for longer antibiotic duration in children with febrile lower respiratory tract infection, and disease severity in adults with SARS-CoV-2 infection.[17,20] |
| **IL-8** | Supportive data from India that increases in IL-8 predict mortality in children with dengue;[19] in Mozambique, IL-8 predicted mortality in children with pneumonia;[14] in the UK, supportive data that increases in IL-8 predict disease severity in children with meningococcal disease.[16] |
| **IL-10** | Supportive data from India that increases in IL-10 predict of mortality in children with dengue.[19] |
| **IP-10** | Supportive data from Uganda that increases in IP-10 predict mortality in children hospitalised with severe malaria.[11] |
| **PCT** | Supportive evidence that increases in PCT predict severe illness in hospitalised children with suspected bacterial infections or meningococcal disease.[21,22] |
| **sTNFR-1** | Supportive data from SSA that increases in sTNFR-1 predict mortality in children hospitalised with pneumonia and all-cause febrile illnesses, and adults with all-cause febrile illnesses.[3,7,8] |
| **sTREM-1** | Supportive data from SSA that increases in sTREM-1 predict mortality in children hospitalised with pneumonia, severe malaria, and all-cause febrile illnesses, and adults with all-cause febrile illnesses;[3,7,8,11,14] in Asia, increased sTREM-1 predicted length of stay in infant febrile illness and in-hospital mortality in adults hospitalised with infection and children hospitalised with pneumonia.[9,23,24] |
| **suPAR** | Supportive data from Uganda that increases in suPAR predict mortality in children with malaria;[25] In Europe, elevated suPAR concentrations predicted length of stay in children with pneumonia and mortality in adults hospitalised with sepsis.[26-28] |

### 6. Table S5: Comparator clinical assessment tools.

| | |
|---|---|
| **WHO Danger Signs (any of the below)** | |
| | Prostration (unable to feed, sit, or stand when previously able) |
| | Intractable vomiting (vomiting everything) |
| | Generalised seizures (witnessed or reported) |
| | Lethargy (abnormally sleepy or AVPU < A) |
| **LqSOFA (score 0-4; one point for each of the below)** | |
| | Abnormal mental status (AVPU < A) |
| | Capillary refill time > 2 seconds |
| | Heart rate > 99$^{th}$ centile for age |
| | Respiratory rate > 99$^{th}$ centile for age |
| **SIRS (score 0-4; one point for each of the below)** | |
| | Core temperature > 38.5 °C or < 36 °C |
| | Heart rate > or < age-adjusted threshold |
| | Respiratory rate > or < age-adjusted threshold |
| | Leukocyte count > or < age-adjusted threshold |

LqSOFA = Liverpool quick Sequential Orga Failure Assessment;[29] SIRS = Systemic Inflammatory Response Syndrome.[30]

### 7. Table S6: Laboratory procedures for biomarker quantification.

Host biomarker concentrations were quantified in EDTA-plasma using the Simple Plex Ella microfluidic platform (ProteinSimple, San Jose, CA, USA) and suPARnostic ELISA (ViroGates, Denmark), or fluoride-oxalate-plasma using LACT2 (Roche Diagnostics, Germany) and GLUC3 (Roche Diagnostics, Germany), according to the manufacturers' protocols. All biomarkers were quantified at the MORU laboratories in Bangkok, Thailand, apart from Indonesian samples, which were quantified at the INA-RESPOND laboratories in Jakarta, Indonesia, with consumables and on-site support provided by the visiting MORU laboratory team.

For the Ella platform, plasma samples were diluted 1:2 (Ang-2, IL-6, IL-8, IL-10, PCT, sTREM-1, sFlt-1), 1:10 (Ang-1, CHI3L1, IL-1ra, IP-10, sTNFR1), or 1:5000 (CRP) in reagent diluent. Analyte concentrations outside the dynamic range of the assay using the initial dilutions were prepared at higher or lower dilutions depending on the raw data relative fluorescent units (RFUs). For suPAR, no dilutions to plasma were performed prior to adding the samples to the pre-coated ELISA plate. Any samples with concentrations above or below the assays' limit of detection were assigned a value of one-third of the highest or lowest limit, respectively, of the limits of quantification. The table below details the proportion of samples assigned a value.

| | **Dynamic Range** (pg/mL) | **Samples outside dynamic range** | **Proportion assigned a value** |
|---|---|---|---|
| Ang-1 | 6.18 - 23,560 | 0 | - |
| Ang-2 | 9.91 - 15,124 | 0 | - |
| CHI3L1 | 6.68 - 25,500 | 2 | 2/3,312 (0.06%) |
| CRP | 32.8 - 50,000 | 5 | 5/3,309 (0.15%) |
| IL-1Ra | 7.37 - 4,500 | 1 | 1/3,313 (0.03%) |
| IL-6 | 0.28 - 2,652 | 4 | 4/3,314 (0.12%) |
| IL-8 | 0.19 - 1,804 | 2 | 2/3,314 (0.06%) |
| IL-10 | 0.58 - 2,212 | 2 | 2/3,314 (0.06%) |
| IP-10 | 0.6 - 920 | 0 | - |
| PCT | 1.58 - 15,100 | 0 | - |
| sFlt-1 | 3.05 - 4,650 | 0 | - |
| sTNFR1 | 0.89 - 3,390 | 0 | - |
| sTREM-1 | 4.2 - 40,000 | 1 | 1/3,314 (0.03%) |
| suPAR | 400 - 16,000* | 15 | - |

*Samples outside the dynamic range (>16,000 pg/mL) for suPAR could be estimated correctly up to 25,000 pg/mL and thus new values were not assigned.

### 8. Figure S1: Study flowchart.

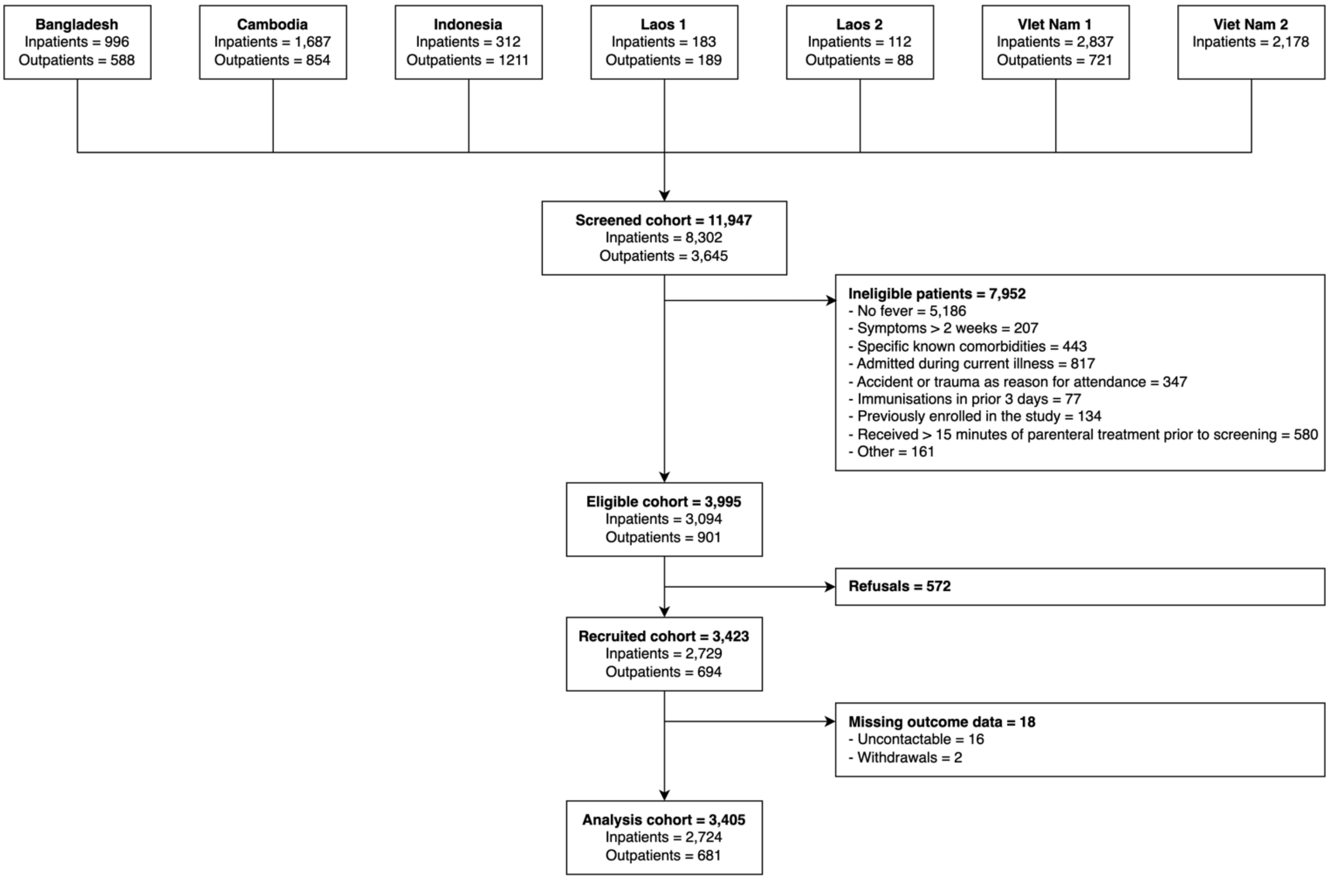

One reason for ineligibility is provided per patient, according to the hierarchy listed in the figure. 1,245 ineligible children had more than one reason for ineligibility (1,245/7,952; 15.7%).

### 9. Table S7: Children receiving care in the community earlier in their illness prior to presentation at the study sites.

| Characteristic | Overall N = 3,405[1] | Bangladesh N = 553[1] | Indonesia N = 187[1] | Cambodia N = 824[1] | Laos 1 N = 207[1] | Laos 2 N = 77[1] | Viet Nam 1 N = 945[1] | Viet Nam 2 N = 612[1] |
|---|---|---|---|---|---|---|---|---|
| **Received care** | 1,342 / 3,405 (39%) | 24 / 553 (4.3%) | 76 / 187 (41%) | 478 / 824 (58%) | 46 / 207 (22%) | 3 / 77 (3.9%) | 496 / 945 (52%) | 219 / 612 (36%) |
| **Location** | | | | | | | | |
| Traditional healer | 6 / 3,405 (0.2%) | 2 / 553 (0.4%) | 0 / 187 (0%) | 1 / 824 (0.1%) | 0 / 207 (0%) | 0 / 77 (0%) | 3 / 945 (0.3%) | 0 / 612 (0%) |
| Private clinic, pharmacy or drug store | 939 / 3,405 (28%) | 11 / 553 (2.0%) | 49 / 187 (26%) | 337 / 824 (41%) | 12 / 207 (5.8%) | 0 / 77 (0%) | 357 / 945 (38%) | 173 / 612 (28%) |
| Primary health centre | 173 / 3,405 (5.1%) | 0 / 553 (0%) | 6 / 187 (3.2%) | 118 / 824 (14%) | 26 / 207 (13%) | 3 / 77 (3.9%) | 10 / 945 (1.1%) | 10 / 612 (1.6%) |
| Hospital outpatient department | 405 / 3,405 (12%) | 11 / 553 (2.0%) | 25 / 187 (13%) | 128 / 824 (16%) | 8 / 207 (3.9%) | 0 / 77 (0%) | 198 / 945 (21%) | 35 / 612 (5.7%) |
| Other | 26 / 3,405 (0.8%) | 0 / 553 (0%) | 14 / 187 (7.5%) | 3 / 824 (0.4%) | 0 / 207 (0%) | 0 / 77 (0%) | 8 / 945 (0.8%) | 1 / 612 (0.2%) |
| **Treatment** | | | | | | | | |
| Antibiotic | 539 / 3,405 (16%) | 11 / 553 (2.0%) | 23 / 187 (12%) | 96 / 824 (12%) | 44 / 207 (21%) | 3 / 77 (3.9%) | 213 / 945 (23%) | 149 / 612 (24%) |
| *Oral* | 518 / 539 (96%) | 11 / 11 (100%) | 23 / 23 (100%) | 93 / 96 (97%) | 36 / 44 (82%) | 0 / 3 (0%) | 206 / 213 (97%) | 149 / 149 (100%) |
| *Parenteral* | 21 / 539 (3.9%) | 0 / 11 (0%) | 0 / 23 (0%) | 3 / 96 (3.1%) | 8 / 44 (18%) | 3 / 3 (100%) | 7 / 213 (3.3%) | 0 / 149 (0%) |
| Intravenous fluid | 61 / 3,405 (1.8%) | 0 / 553 (0%) | 0 / 187 (0%) | 53 / 824 (6.4%) | 0 / 207 (0%) | 1 / 77 (1.3%) | 6 / 945 (0.6%) | 1 / 612 (0.2%) |
| Oxygen/Nebuliser | 94 / 3,405 (2.8%) | 2 / 553 (0.4%) | 4 / 187 (2.1%) | 36 / 824 (4.4%) | 2 / 207 (1.0%) | 1 / 77 (1.3%) | 43 / 945 (4.6%) | 6 / 612 (1.0%) |
| Other/Unknown medication | 1,172 / 3,405 (34%) | 13 / 553 (2.4%) | 61 / 187 (33%) | 439 / 824 (53%) | 4 / 207 (1.9%) | 1 / 77 (1.3%) | 476 / 945 (50%) | 178 / 612 (29%) |
| *Oral* | 1,105 / 1,172 (94%) | 13 / 13 (100%) | 61 / 61 (100%) | 377 / 439 (86%) | 4 / 4 (100%) | 0 / 1 (0%) | 472 / 476 (99%) | 178 / 178 (100%) |
| *Parenteral* | 67 / 1,172 (5.7%) | 0 / 13 (0%) | 0 / 61 (0%) | 62 / 439 (14%) | 0 / 4 (0%) | 1 / 1 (100%) | 4 / 476 (0.8%) | 0 / 178 (0%) |

[1]n / N (%)

### 9. Table S8: Additional presenting clinical characteristics, stratified by whether a child progressed to develop severe disease.

| Characteristic | Overall N = 3,405[1] | Non-severe N = 3,272[1] | Severe N = 133[1] | p-value[2] |
|---|---|---|---|---|
| **Perinatal history** | | | | |
| Birth weight (kg) [*] | 3.1 (2.8, 3.4) | 3.1 (2.8, 3.4) | 3.0 (2.7, 3.3) | 0.091 |
| Low birth weight [a,*] | 325 / 3,366 (9.7%) | 301 / 3,234 (9.3%) | 24 / 132 (18%) | <0.001 |
| Gestation (weeks) [*] | 38.0 (37.0, 39.0) | 38.0 (37.0, 39.0) | 38.0 (37.0, 38.0) | 0.004 |
| Preterm birth [b,*] | 358 / 3,374 (11%) | 338 / 3,242 (10%) | 20 / 132 (15%) | 0.084 |
| Exclusive breastfeeding [*] | 2,382 / 3,382 (70%) | 2,294 / 3,250 (71%) | 88 / 132 (67%) | 0.3 |
| Immunisations up-to-date [*] | 2,871 / 3,381 (85%) | 2,763 / 3,249 (85%) | 108 / 132 (82%) | 0.3 |
| **Anthropometrics** | | | | |
| Weight-for-age z-score (WAZ) | | | | <0.001 |
| *Normal* | 1,890 / 3,405 (56%) | 1,825 / 3,272 (56%) | 65 / 133 (49%) | |
| *1 SD below mean* | 845 / 3,405 (25%) | 821 / 3,272 (25%) | 24 / 133 (18%) | |
| *2 SD below mean* | 399 / 3,405 (12%) | 380 / 3,272 (12%) | 19 / 133 (14%) | |
| *3 SD below mean* | 153 / 3,405 (4.5%) | 142 / 3,272 (4.3%) | 11 / 133 (8.3%) | |
| *4 SD below mean* | 118 / 3,405 (3.5%) | 104 / 3,272 (3.2%) | 14 / 133 (11%) | |
| Weight-for-height z-score (WHZ) [c,*] | | | | <0.001 |
| *Normal* | 2,071 / 3,393 (61%) | 1,994 / 3,261 (61%) | 77 / 132 (58%) | |
| *1 SD below mean* | 737 / 3,393 (22%) | 716 / 3,261 (22%) | 21 / 132 (16%) | |
| *2 SD below mean* | 337 / 3,393 (9.9%) | 326 / 3,261 (10.0%) | 11 / 132 (8.3%) | |
| *3 SD below mean* | 146 / 3,393 (4.3%) | 138 / 3,261 (4.2%) | 8 / 132 (6.1%) | |
| *4 SD below mean* | 102 / 3,393 (3.0%) | 87 / 3,261 (2.7%) | 15 / 132 (11%) | |
| Height-for-age z-score (HAZ) [*] | | | | 0.033 |
| *Normal* | 1,974 / 3,401 (58%) | 1,897 / 3,268 (58%) | 77 / 133 (58%) | |
| *1 SD below mean* | 763 / 3,401 (22%) | 742 / 3,268 (23%) | 21 / 133 (16%) | |
| *2 SD below mean* | 378 / 3,401 (11%) | 363 / 3,268 (11%) | 15 / 133 (11%) | |
| *3 SD below mean* | 165 / 3,401 (4.9%) | 155 / 3,268 (4.7%) | 10 / 133 (7.5%) | |
| *4 SD below mean* | 121 / 3,401 (3.6%) | 111 / 3,268 (3.4%) | 10 / 133 (7.5%) | |
| **Presenting syndrome** | | | | |
| Skin- or soft-tissue infection | 180 / 3,405 (5.3%) | 175 / 3,272 (5.3%) | 5 / 133 (3.8%) | 0.4 |
| Urinary tract infection | 22 / 3,405 (0.6%) | 22 / 3,272 (0.7%) | 0 / 133 (0%) | >0.9 |
| Ear / mastoid | 13 / 3,405 (0.4%) | 12 / 3,272 (0.4%) | 1 / 133 (0.8%) | 0.4 |
| Other | 122 / 3,405 (3.6%) | 106 / 3,272 (3.2%) | 16 / 133 (12%) | <0.001 |
| **Clinical assessment** | | | | |
| Subcostal indrawing | 582 / 3,405 (17%) | 498 / 3,272 (15%) | 84 / 133 (63%) | <0.001 |
| Severe respiratory distress [d] | 339 / 3,405 (10.0%) | 272 / 3,272 (8.3%) | 67 / 133 (50%) | <0.001 |
| Lower limb temperature gradient | 342 / 3,405 (10%) | 321 / 3,272 (9.8%) | 21 / 133 (16%) | 0.025 |

| Characteristic | Overall N = 3,405[1] | Non-severe N = 3,272[1] | Severe N = 133[1] | p-value[2] |
|---|---|---|---|---|
| Reduced skin turgor | 51 / 3,405 (1.5%) | 44 / 3,272 (1.3%) | 7 / 133 (5.3%) | 0.003 |
| Sunken eyes | 306 / 3,405 (9.0%) | 295 / 3,272 (9.0%) | 11 / 133 (8.3%) | 0.8 |
| Ankle oedema | 22 / 3,405 (0.6%) | 18 / 3,272 (0.6%) | 4 / 133 (3.0%) | 0.009 |
| Pallor [e] | 440 / 3,405 (13%) | 399 / 3,272 (12%) | 41 / 133 (31%) | <0.001 |
| Happy child [f] | 658 / 3,405 (19%) | 656 / 3,272 (20%) | 2 / 133 (1.5%) | <0.001 |
| Sepsis at presentation [g] | 670 / 3,405 (20%) | 605 / 3,272 (18%) | 65 / 133 (49%) | <0.001 |
| **Report from caregiver** | | | | |
| Reduced urine output [h,*] | 47 / 3,393 (1.4%) | 41 / 3,261 (1.3%) | 6 / 132 (4.5%) | 0.009 |
| Poor feeding [*] | 1,306 / 3,400 (38%) | 1,222 / 3,267 (37%) | 84 / 133 (63%) | <0.001 |
| Difficulty breathing [*] | 934 / 3,382 (28%) | 838 / 3,250 (26%) | 96 / 132 (73%) | <0.001 |

[1]Median (IQR); n / N (%); [2]Wilcoxon rank sum test; Pearson's Chi-squared test; Fisher's exact test

[a] birthweight < 2.5 kg; [b] gestation < 37 weeks; [c] calculated in children 45-120cm; [d] grunting, tracheal tug, nasal flaring, or intercostal recessions; [e] conjunctival or palmar; [f] assessed by research staff; [g] assessed by treating clinical team; [h] no urine output in last 12 hours.

[*]Missing data: birth weight, n = 513; low birthweight, n = 39; gestation, n = 1,132; preterm birth, n = 31; exclusive breastfeeding, n = 23; immunisations up-to-date, n = 24; WHZ, n = 12; HAZ, n = 4; reduced urine output, n = 12; poor feeding, n = 5; difficulty breathing, n = 23.

### 11. Table S9: Presenting clinical characteristics, stratified by site.

| Characteristic | Overall N = 3,405[1] | Bangladesh N = 553[1] | Indonesia N = 187[1] | Cambodia N = 824[1] | Laos 1 N = 207[1] | Laos 2 N = 77[1] | Viet Nam 1 N = 945[1] | Viet Nam 2 N = 612[1] |
|---|---|---|---|---|---|---|---|---|
| **Demographics and background** | | | | | | | | |
| Age (months) | 16.8 (8.7, 31.0) | 10.5 (5.1, 19.8) | 24.1 (12.5, 35.7) | 13.1 (7.5, 24.4) | 19.9 (11.1, 35.1) | 17.1 (9.0, 34.6) | 20.3 (10.6, 34.3) | 22.9 (12.2, 36.5) |
| Male sex | 2,029 / 3,405 (60%) | 348 / 553 (63%) | 112 / 187 (60%) | 452 / 824 (55%) | 102 / 207 (49%) | 40 / 77 (52%) | 575 / 945 (61%) | 400 / 612 (65%) |
| Comorbidity | 102 / 3,405 (3.0%) | 1 / 553 (0.2%) | 10 / 187 (5.3%) | 37 / 824 (4.5%) | 0 / 207 (0%) | 0 / 77 (0%) | 45 / 945 (4.8%) | 9 / 612 (1.5%) |
| Recent admission | 429 / 3,394 (13%) | 45 / 552 (8.2%) | 16 / 187 (8.6%) | 152 / 820 (19%) | 3 / 207 (1.4%) | 0 / 77 (0%) | 135 / 940 (14%) | 78 / 611 (13%) |
| **Anthropometrics** | | | | | | | | |
| WAZ | -0.8 (-1.7, 0.0) | -2.1 (-3.3, -1.1) | -1.0 (-1.6, -0.1) | -1.2 (-1.9, -0.4) | -1.1 (-2.1, -0.3) | -0.6 (-1.3, 0.2) | -0.3 (-1.0, 0.6) | -0.3 (-1.0, 0.5) |
| Wasted: WHZ < -2* | 585 / 3,393 (17%) | 263 / 550 (48%) | 24 / 187 (13%) | 111 / 820 (14%) | 26 / 207 (13%) | 16 / 77 (21%) | 96 / 941 (10%) | 49 / 611 (8.0%) |
| Stunted: HAZ < -2* | 664 / 3,401 (20%) | 180 / 553 (33%) | 33 / 187 (18%) | 180 / 822 (22%) | 86 / 207 (42%) | 5 / 77 (6.5%) | 111 / 943 (12%) | 69 / 612 (11%) |
| **Illness history** | | | | | | | | |
| Illness days | 3.0 (2.0, 4.0) | 3.0 (2.0, 5.0) | 3.0 (1.0, 4.0) | 3.0 (2.0, 4.0) | 3.0 (2.0, 4.0) | 3.0 (2.0, 5.0) | 3.0 (2.0, 4.0) | 3.0 (2.0, 4.0) |
| Sought care prior | 1,753 / 3,405 (51%) | 40 / 553 (7.2%) | 161 / 187 (86%) | 641 / 824 (78%) | 57 / 207 (28%) | 4 / 77 (5.2%) | 600 / 945 (63%) | 250 / 612 (41%) |
| Travel time ≤ 1 h | 2,777 / 3,405 (82%) | 539 / 553 (97%) | 175 / 187 (94%) | 550 / 824 (67%) | 183 / 207 (88%) | 73 / 77 (95%) | 840 / 945 (89%) | 417 / 612 (68%) |
| **Presenting syndrome** | | | | | | | | |
| URTI | 1,121 / 3,405 (33%) | 130 / 553 (24%) | 59 / 187 (32%) | 326 / 824 (40%) | 174 / 207 (84%) | 41 / 77 (53%) | 338 / 945 (36%) | 53 / 612 (8.7%) |
| LRTI | 1,347 / 3,405 (40%) | 189 / 553 (34%) | 69 / 187 (37%) | 417 / 824 (51%) | 48 / 207 (23%) | 26 / 77 (34%) | 384 / 945 (41%) | 214 / 612 (35%) |
| Diarrhoeal | 646 / 3,405 (19%) | 102 / 553 (18%) | 34 / 187 (18%) | 249 / 824 (30%) | 13 / 207 (6.3%) | 6 / 77 (7.8%) | 165 / 945 (17%) | 77 / 612 (13%) |
| Neurological | 430 / 3,405 (13%) | 60 / 553 (11%) | 28 / 187 (15%) | 58 / 824 (7.0%) | 1 / 207 (0.5%) | 3 / 77 (3.9%) | 207 / 945 (22%) | 73 / 612 (12%) |
| No focus | 527 / 3,405 (15%) | 39 / 553 (7.1%) | 41 / 187 (22%) | 155 / 824 (19%) | 1 / 207 (0.5%) | 4 / 77 (5.2%) | 222 / 945 (23%) | 65 / 612 (11%) |
| **Severity at presentation** | | | | | | | | |
| WHO Danger Sign* | 1,607 / 3,398 (47%) | 233 / 552 (42%) | 101 / 187 (54%) | 361 / 823 (44%) | 81 / 207 (39%) | 13 / 75 (17%) | 501 / 942 (53%) | 317 / 612 (52%) |

| Characteristic | Overall N = 3,405[1] | Bangladesh N = 553[1] | Indonesia N = 187[1] | Cambodia N = 824[1] | Laos 1 N = 207[1] | Laos 2 N = 77[1] | Viet Nam 1 N = 945[1] | Viet Nam 2 N = 612[1] |
|---|---|---|---|---|---|---|---|---|
| *Prostration* | 240 / 3,405 (7.0%) | 121 / 553 (22%) | 13 / 187 (7.0%) | 41 / 824 (5.0%) | 2 / 207 (1.0%) | 1 / 77 (1.3%) | 18 / 945 (1.9%) | 44 / 612 (7.2%) |
| *Vomiting** | 687 / 3,401 (20%) | 101 / 553 (18%) | 29 / 186 (16%) | 83 / 823 (10%) | 51 / 207 (25%) | 3 / 76 (3.9%) | 204 / 944 (22%) | 216 / 612 (35%) |
| *Seizures** | 433 / 3,400 (13%) | 62 / 551 (11%) | 31 / 187 (17%) | 60 / 823 (7.3%) | 4 / 206 (1.9%) | 1 / 76 (1.3%) | 211 / 945 (22%) | 64 / 612 (10%) |
| *Lethargy** | 825 / 3,399 (24%) | 86 / 551 (16%) | 62 / 187 (33%) | 305 / 824 (37%) | 35 / 207 (17%) | 11 / 76 (14%) | 212 / 943 (22%) | 114 / 611 (19%) |
| LqSOFA score* | | | | | | | | |
| *0* | 2,639 / 3,402 (78%) | 319 / 553 (58%) | 166 / 187 (89%) | 641 / 823 (78%) | 189 / 207 (91%) | 70 / 77 (91%) | 713 / 943 (76%) | 541 / 612 (88%) |
| *1* | 642 / 3,402 (19%) | 167 / 553 (30%) | 19 / 187 (10%) | 156 / 823 (19%) | 18 / 207 (8.7%) | 5 / 77 (6.5%) | 213 / 943 (23%) | 64 / 612 (10%) |
| *2* | 98 / 3,402 (2.9%) | 49 / 553 (8.9%) | 1 / 187 (0.5%) | 23 / 823 (2.8%) | 0 / 207 (0%) | 1 / 77 (1.3%) | 17 / 943 (1.8%) | 7 / 612 (1.1%) |
| *3* | 20 / 3,402 (0.6%) | 15 / 553 (2.7%) | 1 / 187 (0.5%) | 3 / 823 (0.4%) | 0 / 207 (0%) | 1 / 77 (1.3%) | 0 / 943 (0%) | 0 / 612 (0%) |
| *4* | 3 / 3,402 (<0.1%) | 3 / 553 (0.5%) | 0 / 187 (0%) | 0 / 823 (0%) | 0 / 207 (0%) | 0 / 77 (0%) | 0 / 943 (0%) | 0 / 612 (0%) |
| SIRS score* | | | | | | | | |
| *0* | 223 / 2,827 (7.9%) | 1 / 180 (0.6%) | 19 / 171 (11%) | 70 / 814 (8.6%) | 24 / 99 (24%) | 4 / 46 (8.7%) | 48 / 908 (5.3%) | 57 / 609 (9.4%) |
| *1* | 1,042 / 2,827 (37%) | 60 / 180 (33%) | 59 / 171 (35%) | 378 / 814 (46%) | 39 / 99 (39%) | 13 / 46 (28%) | 297 / 908 (33%) | 196 / 609 (32%) |
| *2* | 916 / 2,827 (32%) | 71 / 180 (39%) | 56 / 171 (33%) | 230 / 814 (28%) | 27 / 99 (27%) | 21 / 46 (46%) | 302 / 908 (33%) | 209 / 609 (34%) |
| *3* | 495 / 2,827 (18%) | 35 / 180 (19%) | 32 / 171 (19%) | 113 / 814 (14%) | 6 / 99 (6.1%) | 6 / 46 (13%) | 197 / 908 (22%) | 106 / 609 (17%) |
| *4* | 151 / 2,827 (5.3%) | 13 / 180 (7.2%) | 5 / 171 (2.9%) | 23 / 814 (2.8%) | 3 / 99 (3.0%) | 2 / 46 (4.3%) | 64 / 908 (7.0%) | 41 / 609 (6.7%) |
| **Vital signs** | | | | | | | | |
| Heart rate* | | | | | | | | |
| *1-12m (bpm)* | 156.0 (140.0, 172.0) | 151.0 (140.0, 170.0) | 150.0 (142.0, 161.0) | 159.0 (143.0, 175.5) | 149.0 (131.0, 162.0) | 125.0 (114.0, 126.0) | 165.0 (147.0, 178.2) | 149.0 (135.0, 169.0) |
| *12-60m (bpm)* | 140.0 (127.0, 158.0) | 140.0 (128.0, 153.5) | 136.0 (123.2, 148.8) | 140.0 (128.0, 162.0) | 136.0 (113.0, 150.0) | 112.0 (106.0, 122.0) | 148.0 (133.0, 165.0) | 138.0 (125.0, 150.0) |
| Respiratory rate* | | | | | | | | |
| *1-12m (bpm)* | 45.0 (38.0, 55.0) | 48.0 (39.0, 60.0) | 39.0 (30.0, 50.0) | 44.0 (37.0, 54.0) | 32.0 (29.5, 42.5) | 43.0 (42.0, 46.8) | 46.0 (40.0, 52.0) | 40.0 (35.0, 50.0) |
| *12-60m (bpm)* | 36.0 (30.0, 42.0) | 38.0 (34.0, 44.0) | 31.5 (26.0, 39.0) | 38.0 (32.0, 45.0) | 29.0 (26.0, 32.0) | 32.0 (30.0, 36.5) | 38.0 (32.0, 44.0) | 32.0 (30.0, 39.0) |
| $SpO_2$ (%)* | 98.0 (97.0, 99.0) | 99.0 (98.0, 100.0) | 99.0 (98.0, 100.0) | 99.0 (98.0, 100.0) | 99.0 (98.0, 100.0) | 100.0 (99.0, 100.0) | 97.0 (96.0, 99.0) | 98.0 (96.0, 98.0) |

| Characteristic | Overall N = 3,405[1] | Bangladesh N = 553[1] | Indonesia N = 187[1] | Cambodia N = 824[1] | Laos 1 N = 207[1] | Laos 2 N = 77[1] | Viet Nam 1 N = 945[1] | Viet Nam 2 N = 612[1] |
|---|---|---|---|---|---|---|---|---|
| Temperature (°C)* | 37.6 (37.0, 38.3) | 37.6 (37.0, 38.3) | 38.0 (37.3, 38.8) | 37.3 (36.7, 38.0) | 36.8 (36.2, 37.6) | 37.7 (37.5, 38.3) | 37.8 (37.2, 38.5) | 37.8 (37.0, 38.5) |
| Prolonged CRT | 194 / 3,405 (5.7%) | 173 / 553 (31%) | 1 / 187 (0.5%) | 14 / 824 (1.7%) | 0 / 207 (0%) | 2 / 77 (2.6%) | 1 / 945 (0.1%) | 3 / 612 (0.5%) |
| Not alert | 86 / 3,405 (2.5%) | 34 / 553 (6.1%) | 7 / 187 (3.7%) | 28 / 824 (3.4%) | 0 / 207 (0%) | 3 / 77 (3.9%) | 12 / 945 (1.3%) | 2 / 612 (0.3%) |
| **Biomarkers** | | | | | | | | |
| Endothelial activation | | | | | | | | |
| *Ang-1 (pg/ml)** | 6,156.0 (3,407.5, 10,833.2) | 7,992.0 (4,093.5, 15,372.5) | 4,377.5 (2,256.0, 8,547.0) | 5,686.5 (3,454.5, 9,134.0) | 8,502.0 (5,260.5, 11,794.0) | 7,875.5 (5,309.2, 10,528.8) | 5,973.0 (3,186.0, 10,427.0) | 5,719.0 (3,261.2, 10,553.0) |
| *Ang-2 (pg/ml)** | 1,359.0 (1,021.2, 1,883.0) | 1,755.0 (1,258.5, 2,736.0) | 1,104.5 (890.0, 1,518.0) | 1,455.5 (1,090.2, 2,050.2) | 1,229.5 (1,014.8, 1,558.0) | 1,266.0 (903.2, 1,736.5) | 1,265.0 (964.0, 1,720.0) | 1,276.0 (971.5, 1,666.2) |
| *sFlt-1 (pg/ml)** | 186.0 (150.0, 235.0) | 187.0 (134.5, 246.5) | 164.0 (138.5, 194.0) | 196.0 (156.0, 245.0) | 178.0 (150.8, 219.2) | 173.5 (146.2, 230.2) | 186.0 (151.0, 231.0) | 186.0 (154.8, 229.0) |
| Immune activation | | | | | | | | |
| *CHI3L1 (ng/ml)** | 25.0 (16.7, 39.1) | 36.5 (24.0, 55.1) | 15.4 (11.7, 22.8) | 28.9 (20.2, 43.1) | 26.9 (19.2, 42.5) | 28.0 (19.4, 40.9) | 22.1 (15.1, 33.2) | 20.1 (13.6, 30.4) |
| *CRP (mg/l)** | 14.3 (3.5, 51.4) | 10.7 (3.1, 41.4) | 4.7 (1.4, 19.2) | 17.3 (4.8, 55.2) | 11.7 (2.8, 36.4) | 16.2 (6.0, 69.5) | 9.8 (2.4, 38.2) | 27.7 (7.7, 110.0) |
| *IL-1ra (pg/ml)** | 1,966.0 (920.0, 4,795.0) | 1,647.0 (694.0, 4,555.0) | 1,728.5 (1,046.2, 3,977.0) | 1,755.0 (790.5, 4,425.8) | 1,240.0 (586.0, 2,707.0) | 1,652.0 (750.5, 2,879.8) | 2,474.0 (1,205.5, 5,934.0) | 2,125.0 (1,094.0, 4,485.0) |
| *IL-6 (pg/ml)** | 17.9 (7.6, 46.3) | 16.9 (6.6, 42.6) | 13.9 (6.5, 37.5) | 16.9 (7.2, 46.9) | 15.1 (6.6, 34.5) | 21.9 (8.9, 43.5) | 18.3 (7.7, 45.6) | 21.4 (8.9, 59.5) |
| *IL-8 (pg/ml)** | 15.4 (9.2, 27.5) | 23.5 (13.6, 44.2) | 19.1 (12.4, 28.5) | 15.6 (9.3, 27.5) | 13.2 (8.7, 20.6) | 10.5 (6.8, 19.0) | 14.4 (8.9, 26.6) | 12.2 (7.6, 21.2) |
| *IL-10 (pg/ml)** | 19.5 (10.5, 43.6) | 19.7 (11.4, 35.8) | 22.7 (12.0, 47.4) | 20.4 (10.8, 43.6) | 14.1 (9.1, 30.4) | 17.8 (9.1, 40.9) | 21.6 (10.9, 55.8) | 17.4 (9.2, 41.2) |
| *IP-10 (pg/ml)** | 825.0 (381.0, 1,878.0) | 583.0 (272.5, 1,090.5) | 875.0 (373.0, 1,846.5) | 727.0 (368.0, 1,857.5) | 710.5 (366.5, 1,317.8) | 833.0 (418.2, 1,408.2) | 1,320.0 (518.5, 2,969.0) | 791.5 (375.5, 1,583.2) |
| *PCT (ng/ml)** | 0.3 (0.2, 0.8) | 0.4 (0.3, 1.0) | 0.2 (0.1, 0.4) | 0.4 (0.2, 1.1) | 0.3 (0.2, 0.8) | 0.4 (0.2, 0.8) | 0.3 (0.2, 0.7) | 0.3 (0.2, 0.8) |
| *sTNFR-1 (pg/ml)** | 1,581.5 (1,257.0, 2,055.0) | 1,688.0 (1,309.5, 2,320.0) | 1,335.0 (1,152.2, 1,696.0) | 1,643.5 (1,306.2, 2,153.0) | 1,378.0 (1,175.0, 1,748.8) | 1,568.5 (1,287.5, 1,843.5) | 1,636.0 (1,335.0, 2,093.5) | 1,484.5 (1,180.8, 1,882.2) |
| *sTREM-1 (pg/ml)** | 230.0 (169.0, 332.0) | 323.0 (232.0, 454.0) | 204.5 (153.0, 272.8) | 238.0 (182.2, 330.8) | 215.5 (175.0, 279.2) | 212.0 (163.0, 331.0) | 196.0 (145.0, 275.0) | 227.5 (171.0, 320.2) |

| Characteristic | Overall<br>N = 3,405[1] | Bangladesh<br>N = 553[1] | Indonesia<br>N = 187[1] | Cambodia<br>N = 824[1] | Laos 1<br>N = 207[1] | Laos 2<br>N = 77[1] | Viet Nam 1<br>N = 945[1] | Viet Nam 2<br>N = 612[1] |
|---|---|---|---|---|---|---|---|---|
| *suPAR (ng/ml)* * | 4.2<br>(3.4, 5.2) | 4.8<br>(3.9, 5.9) | 3.8<br>(3.3, 4.5) | 4.6<br>(3.7, 5.8) | 3.8<br>(3.3, 4.7) | 3.9<br>(3.4, 4.7) | 4.0<br>(3.2, 4.9) | 3.7<br>(3.0, 4.7) |
| Other | | | | | | | | |
| *Lactate (mmol/l)* * | 1.0<br>(0.8, 1.4) | 1.0<br>(0.7, 1.4) | 1.0<br>(0.7, 1.3) | 1.2<br>(0.9, 1.6) | 1.1<br>(0.8, 1.5) | 1.2<br>(1.0, 1.6) | 1.0<br>(0.7, 1.3) | 0.9<br>(0.7, 1.2) |
| *Glucose (mmol/l)* * | 5.4<br>(4.8, 6.2) | 5.6<br>(4.9, 6.4) | 5.1<br>(4.6, 6.0) | 5.5<br>(4.9, 6.4) | 5.3<br>(4.7, 5.8) | 5.2<br>(4.7, 5.7) | 5.4<br>(4.9, 6.2) | 5.3<br>(4.8, 6.1) |
| *Hb (g/dL)* * | 11.3<br>(10.3, 12.2) | 10.2<br>(9.3, 11.2) | 11.6<br>(10.9, 12.2) | 10.8<br>(9.9, 11.6) | 11.7<br>(10.5, 12.3) | 11.2<br>(10.4, 11.9) | 11.4<br>(10.5, 12.3) | 11.9<br>(11.1, 12.6) |

[1]Median (IQR); n / N (%)

*Missing data: weight-for-height z-score (WHZ), n = 12; height-for-age z-score (HAZ), n = 4; WHO Danger Sign, n = 7; vomiting everything, n = 4; generalised seizures, n = 5; lethargy, n = 6; LqSOFA, n = 3; SIRS, n = 578; heart rate, n = 1; respiratory rate, n = 2; oxygen saturation, n = 205; axillary temperature, n = 1; Ang-1, Ang-2, sFlt-1, IL-6, IL-8, IL-10, IP-10, PCT, sTNFR-1, sTREM-1, n = 91; CHI3L1, n = 93; CRP, n = 96; IL-1ra, n = 92; suPAR, n = 124; lactate, glucose, n = 98; Hb, n = 574. Missingness for SIRS and Hb was greater, as complete blood counts were measured at the discretion of the treating clinical team.

### 12. Table S10: Presenting concentrations of immune and endothelial activation markers, stratified by whether a child progressed to develop severe disease.

| Characteristic | Overall<br>N = 3,405[1] | Non-severe<br>N = 3,272[1] | Severe<br>N = 133[1] | p-value[2] |
|---|---|---|---|---|
| **Endothelial activation** | | | | |
| Ang-1 (pg/ml) | 6,156.0 (3,407.5, 10,833.2) | 6,186.0 (3,404.5, 10,834.0) | 5,746.0 (3,443.5, 10,456.0) | 0.5 |
| Ang-2 (pg/ml) | 1,359.0 (1,021.2, 1,883.0) | 1,343.0 (1,014.0, 1,845.5) | 2,532.0 (1,445.5, 4,318.5) | <0.001 |
| sFlt-1 (pg/ml) | 186.0 (150.0, 235.0) | 184.0 (149.0, 230.0) | 258.0 (204.0, 370.0) | <0.001 |
| **Immune activation** | | | | |
| CHI3L1 (ng/ml) | 25.0 (16.7, 39.1) | 24.5 (16.6, 38.3) | 37.8 (24.3, 66.2) | <0.001 |
| CRP (mg/l) | 14.3 (3.5, 51.4) | 14.0 (3.5, 49.9) | 25.8 (5.2, 103.2) | <0.001 |
| IL-1ra (pg/ml) | 1,966.0 (920.0, 4,795.0) | 1,932.0 (914.0, 4,675.2) | 2,829.0 (1,219.0, 9,712.5) | <0.001 |
| IL-6 (pg/ml) | 17.9 (7.6, 46.3) | 17.5 (7.5, 43.9) | 51.8 (11.5, 197.0) | <0.001 |
| IL-8 (pg/ml) | 15.4 (9.2, 27.5) | 15.1 (9.1, 26.7) | 28.7 (14.3, 85.3) | <0.001 |
| IL-10 (pg/ml) | 19.5 (10.5, 43.6) | 19.2 (10.5, 42.7) | 27.5 (14.2, 69.1) | <0.001 |
| IP-10 (pg/ml) | 825.0 (381.0, 1,878.0) | 836.0 (390.5, 1,898.0) | 591.0 (234.0, 1,209.5) | <0.001 |
| PCT (ng/ml) | 0.3 (0.2, 0.8) | 0.3 (0.2, 0.8) | 0.8 (0.4, 3.5) | <0.001 |
| sTNFR-1 (pg/ml) | 1,581.5 (1,257.0, 2,055.0) | 1,569.0 (1,249.0, 2,032.5) | 2,121.0 (1,550.0, 3,166.5) | <0.001 |
| sTREM-1 (pg/ml) | 230.0 (169.0, 332.0) | 227.0 (167.0, 323.0) | 376.0 (273.0, 563.5) | <0.001 |
| suPAR (ng/ml) | 4.2 (3.4, 5.2) | 4.1 (3.3, 5.2) | 5.4 (4.1, 7.7) | <0.001 |
| **Other** | | | | |
| Lactate (mmol/l) | 1.0 (0.8, 1.4) | 1.0 (0.7, 1.4) | 1.4 (0.8, 2.1) | <0.001 |
| Glucose (mmol/l) | 5.4 (4.8, 6.2) | 5.4 (4.8, 6.2) | 6.0 (5.1, 7.1) | <0.001 |
| Hb (g/dL) | 11.3 (10.3, 12.2) | 11.3 (10.4, 12.2) | 10.8 (9.5, 12.0) | <0.001 |

[1]Median (IQR); [2]Wilcoxon rank sum test

Missing data: Ang-1, Ang-2, sFlt-1, IL-6, IL-8, IL-10, IP-10, PCT, sTNFR-1, sTREM-1, n = 91; CHI3L1, n = 93; CRP, n = 96; IL-1ra, n = 92; suPAR, n = 124; lactate, glucose, n = 98; Hb, n = 574; WHO Danger Sign, n = 7, LqSOFA, n = 3, SIRS, n = 578. Missingness for SIRS and Hb was greater, as complete blood counts were measured at the discretion of the treating clinical team.

### 13. Table S11: Identified causes of acute febrile illness.

| Pathogen | Overall N = 3,405[1] | Bangladesh N = 553[1] | Cambodia N = 824[1] | Indonesia N = 187[1] | Laos 1 N = 207[1] | Laos 2 N = 77[1] | Viet Nam 1 N = 945[1] | Viet Nam 2 N = 612[1] |
|---|---|---|---|---|---|---|---|---|
| Zika | 8 / 3,289 (0.2%) | 0 / 517 (0%) | 7 / 815 (0.9%) | 0 / 161 (0%) | 0 / 197 (0%) | 0 / 72 (0%) | 1 / 933 (0.1%) | 0 / 594 (0%) |
| Dengue | 109 / 3,289 (3.3%) | 1 / 517 (0.2%) | 2 / 815 (0.2%) | 3 / 161 (1.9%) | 5 / 197 (2.5%) | 0 / 72 (0%) | 97 / 933 (10%) | 1 / 594 (0.2%) |
| Chikungunya | 47 / 3,289 (1.4%) | 0 / 517 (0%) | 43 / 815 (5.3%) | 0 / 161 (0%) | 2 / 197 (1.0%) | 0 / 72 (0%) | 0 / 933 (0%) | 2 / 594 (0.3%) |
| *Leptospirosis* spp. | 4 / 3,128 (0.1%) | 0 / 517 (0%) | 0 / 815 (0%) | - | 1 / 197 (0.5%) | 1 / 72 (1.4%) | 1 / 933 (0.1%) | 1 / 594 (0.2%) |
| *Rickettsia* spp. | 6 / 3,128 (0.2%) | 3 / 517 (0.6%) | 1 / 815 (0.1%) | - | 0 / 197 (0%) | 1 / 72 (1.4%) | 0 / 933 (0%) | 1 / 594 (0.2%) |
| *Orientia tsutsugamushi* | 3 / 3,128 (<0.1%) | 0 / 517 (0%) | 2 / 815 (0.2%) | - | 0 / 197 (0%) | 0 / 72 (0%) | 1 / 933 (0.1%) | 0 / 594 (0%) |
| SARS-CoV-2 | 81 / 2,861 (2.8%) | 15 / 507 (3.0%) | 7 / 652 (1.1%) | - | 0 / 193 (0%) | 1 / 77 (1.3%) | 37 / 842 (4.4%) | 21 / 590 (3.6%) |
| Influenza A | 87 / 2,861 (3.0%) | 28 / 507 (5.5%) | 18 / 652 (2.8%) | - | 2 / 193 (1.0%) | 4 / 77 (5.2%) | 18 / 842 (2.1%) | 17 / 590 (2.9%) |
| Influenza B | 59 / 2,861 (2.1%) | 25 / 507 (4.9%) | 0 / 652 (0%) | - | 0 / 193 (0%) | 0 / 77 (0%) | 29 / 842 (3.4%) | 5 / 590 (0.8%) |
| Respiratory syncytial virus | 429 / 2,861 (15%) | 55 / 507 (11%) | 80 / 652 (12%) | - | 58 / 193 (30%) | 0 / 77 (0%) | 113 / 842 (13%) | 123 / 590 (21%) |
| Human metapneumovirus | 65 / 2,209 (2.9%) | 39 / 507 (7.7%) | - | - | 1 / 193 (0.5%) | 0 / 77 (0%) | 19 / 842 (2.3%) | 6 / 590 (1.0%) |
| *Mycoplasma pneumoniae* | 3 / 2,209 (0.1%) | 0 / 507 (0%) | - | - | 0 / 193 (0%) | 0 / 77 (0%) | 3 / 842 (0.4%) | 0 / 590 (0%) |
| *Chlamydia pneumoniae* | 3 / 2,209 (0.1%) | 2 / 507 (0.4%) | - | - | 1 / 193 (0.5%) | 0 / 77 (0%) | 0 / 842 (0%) | 0 / 590 (0%) |
| *Bordetella parapertussis* | 8 / 2,209 (0.4%) | 5 / 507 (1.0%) | - | - | 2 / 193 (1.0%) | 1 / 77 (1.3%) | 0 / 842 (0%) | 0 / 590 (0%) |
| *Bordetella pertussis* | 1 / 2,209 (<0.1%) | 1 / 507 (0.2%) | - | - | 0 / 193 (0%) | 0 / 77 (0%) | 0 / 842 (0%) | 0 / 590 (0%) |
| Bacteraemia* | 19 / 1,192 (1.6%) | 1 / 194 (0.5%) | 7 / 411 (1.7%) | 1 / 9 (11%) | 0 / 6 (0%) | 1 / 46 (2.2%) | 8 / 386 (2.1%) | 1 / 140 (0.7%) |

[1]n / N (%)

*Bacteraemia diagnosed by blood culture: *Staphylococcus aureus*, n = 5; *Escherichia coli*, n = 3; *Acinetobacter* spp., n = 2; *Salmonella* spp., n = 2; Other, n = 7 (*Campylobacter jejuni*, n = 1; *Enterobacter* spp., n = 1; *Enterococcus faecalis*, n = 1; *Haemophilus influenzae*, n = 1; *Streptococcus pneumoniae*, n = 1; *Streptococcus pyogenes*, n = 1; *Vibrio* spp., n = 1).

In Cambodia, respiratory samples were tested for SARS-CoV-2, influenza A, influenza B, and RSV. In Indonesia, PCR testing was performed for Zika, dengue, and chikungunya. When feasible, the study supported collection of blood cultures at the discretion of the treating clinical team.

14. **Table S12: Prognostic performance of endothelial and immune activation markers and clinical assessment tools to predict progression to severe disease within two days of enrolment in children with microbiologically-confirmed infections, WHO-pneumonia, and non-respiratory presentations.**

| | AUC (95% CI)<br>N = events / non-events | | | |
|---|---|---|---|---|
| | **Overall**<br>N = 133 / 3,272 | **Microbiologically-confirmed infections**<br>N = 49 / 849 | **WHO-pneumonia**<br>N = 85 / 1,024 | **Non-respiratory presentations**<br>N = 44 / 1,527 |
| **Clinical assessment tools** | | | | |
| WHO Danger Sign | 0.75 (0.71-0.80) | 0.76 (0.69-0.83) | 0.58 (0.50-0.65) | 0.83 (0.77-0.89) |
| LqSOFA | 0.74 (0.69-0.78) | 0.73 (0.65-0.80) | 0.72 (0.66-0.78) | 0.67 (0.60-0.76) |
| SIRS | 0.63 (0.58-0.68) | 0.51 (0.41-0.61) | 0.62 (0.53-0.70) | 0.64 (0.55-0.73) |
| **Biomarkers** | | | | |
| sTREM-1 | 0.86 (0.82-0.90) | 0.88 (0.83-0.94) | 0.84 (0.78-0.89) | 0.87 (0.81-0.94) |
| sFlt-1 | 0.81 (0.77-0.86) | 0.82 (0.75-0.89) | 0.84 (0.77-0.90) | 0.73 (0.64-0.83) |
| Ang-2 | 0.80 (0.75-0.85) | 0.83 (0.76-0.90) | 0.85 (0.79-0.91) | 0.69 (0.58-0.79) |
| PCT | 0.80 (0.75-0.84) | 0.77 (0.70-0.85) | 0.78 (0.71-0.85) | 0.81 (0.72-0.89) |
| sTNFR-1 | 0.77 (0.72-0.82) | 0.79 (0.72-0.86) | 0.78 (0.71-0.85) | 0.78 (0.69-0.86) |
| IL-8 | 0.76 (0.70-0.81) | 0.80 (0.72-0.88) | 0.74 (0.66-0.82) | 0.72 (0.61-0.83) |
| IL-6 | 0.74 (0.69-0.79) | 0.69 (0.60-0.78) | 0.67 (0.58-0.76) | 0.85 (0.78-0.93) |
| CHI3L1 | 0.74 (0.68-0.79) | 0.69 (0.60-0.78) | 0.77 (0.69-0.84) | 0.75 (0.66-0.84) |
| suPAR | 0.69 (0.64-0.75) | 0.71 (0.63-0.80) | 0.74 (0.66-0.82) | 0.60 (0.48-0.72) |
| Lactate | 0.68 (0.62-0.74) | 0.69 (0.59-0.78) | 0.71 (0.63-0.79) | 0.57 (0.45-0.69) |
| Glucose | 0.67 (0.61-0.73) | 0.64 (0.54-0.74) | 0.64 (0.55-0.72) | 0.63 (0.52-0.73) |
| IL-1ra | 0.66 (0.60-0.72) | 0.57 (0.48-0.67) | 0.65 (0.55-0.74) | 0.71 (0.63-0.81) |
| CRP | 0.64 (0.58-0.70) | 0.68 (0.58-0.77) | 0.59 (0.50-0.68) | 0.73 (0.64-0.82) |
| IP-10* | 0.64 (0.58-0.69) | 0.69 (0.62-0.77) | 0.59 (0.49-0.68) | 0.65 (0.55-0.75) |
| IL-10 | 0.63 (0.58-0.69) | 0.60 (0.50-0.69) | 0.57 (0.48-0.67) | 0.66 (0.56-0.75) |
| Haemoglobin* | 0.60 (0.54-0.67) | 0.70 (0.60-0.80) | 0.70 (0.62-0.79) | 0.49 (0.37-0.61) |
| Ang-1* | 0.48 (0.42-0.53) | 0.42 (0.33-0.51) | 0.53 (0.43-0.62) | 0.48 (0.38-0.57) |

*Reciprocal concentration used, as lower biomarker concentrations known to be indicative of more severe disease.

15. **Table S13: Site-specific prognostic performance of endothelial and immune activation markers and clinical assessment tools to predict progression to severe disease within two days of enrolment.**

| | **AUC (95% CI)** N = events / non-events | | | | | |
|---|---|---|---|---|---|---|
| | **Overall** N = 133 / 3,272 | **Bangladesh** N = 39 / 514 | **Cambodia** N = 36 / 788 | **Viet Nam 1** N = 32 / 913 | **Viet Nam 2** N = 26 / 586 | **Excluding Viet Nam 2** N = 107 / 2,686 |
| **Clinical assessment tools** | | | | | | |
| WHO Danger Sign | 0.75 (0.71-0.80) | 0.80 (0.73-0.87) | 0.69 (0.61-0.78) | 0.77 (0.69-0.85) | 0.75 (0.66-0.84) | 0.75 (0.71-0.80) |
| LqSOFA | 0.74 (0.69-0.78) | 0.82 (0.75-0.90) | 0.63 (0.55-0.72) | 0.84 (0.76-0.92) | 0.60 (0.51-0.69) | 0.77 (0.72-0.82) |
| SIRS | 0.63 (0.58-0.68) | 0.53 (0.38-0.69) | 0.60 (0.51-0.68) | 0.64 (0.54-0.75) | 0.69 (0.58-0.81) | 0.620.58-0.66) |
| **Biomarkers** | | | | | | |
| sTREM-1 | 0.86 (0.82-0.90) | 0.87 (0.80-0.93) | 0.84 (0.76-0.92) | 0.87 (0.79-0.95) | 0.89 (0.82-0.96) | 0.86 (0.81-0.90) |
| sFlt-1 | 0.81 (0.77-0.86) | 0.88 (0.78-0.97) | 0.87 (0.81-0.92) | 0.77 (0.67-0.86) | 0.71 (0.59-0.81) | 0.84 (0.79-0.89) |
| Ang-2 | 0.80 (0.75-0.85) | 0.89 (0.83-0.96) | 0.85 (0.76-0.94) | 0.79 (0.69-0.89) | 0.63 (0.50-0.75) | 0.84 (0.79-0.89) |
| PCT | 0.80 (0.75-0.84) | 0.82 (0.75-0.89) | 0.75 (0.67-0.82) | 0.79 (0.69-0.88) | 0.81 (0.72-0.91) | 0.79 (0.75-0.84) |
| sTNFR-1 | 0.77 (0.72-0.82) | 0.86 (0.79-0.93) | 0.78 (0.69-0.86) | 0.67 (0.56-0.78) | 0.76 (0.66-0.87) | 0.77 (0.72-0.83) |
| IL-8 | 0.76 (0.70-0.81) | 0.88 (0.81-0.95) | 0.77 (0.68-0.86) | 0.76 (0.65-0.87) | 0.60 (0.46-0.73) | 0.79 (0.74-0.85) |
| IL-6 | 0.74 (0.69-0.79) | 0.77 (0.66-0.87) | 0.60 (0.48-0.71) | 0.77 (0.67-0.86) | 0.87 (0.79-0.95) | 0.71 (0.64-0.77) |
| CHI3L1 | 0.74 (0.68-0.79) | 0.70 (0.59-0.79) | 0.71 (0.62-0.81) | 0.72 (0.62-0.83) | 0.82 (0.71-0.93) | 0.71 (0.65-0.76) |
| suPAR | 0.69 (0.64-0.75) | 0.77 (0.68-0.87) | 0.73 (0.65-0.82) | 0.70 (0.59-0.80) | 0.47 (0.33-0.61) | 0.74 (0.69-0.79) |
| Lactate | 0.68 (0.62-0.74) | 0.82 (0.74-0.91) | 0.83 (0.75-0.91) | 0.56 (0.44-0.67) | 0.51 (0.38-0.64) | 0.73 (0.67-0.79) |
| Glucose | 0.67 (0.61-0.73) | 0.71 (0.59-0.83) | 0.61 (0.50-0.73) | 0.71 (0.61-0.82) | 0.63 (0.50-0.76) | 0.68 (0.61-0.74) |
| IL-1ra | 0.66 (0.60-0.72) | 0.76 (0.66-0.86) | 0.55 (0.45-0.65) | 0.66 (0.56-0.77) | 0.68 (0.56-0.79) | 0.65 (0.59-0.72) |

| | | | | | | |
|---|---|---|---|---|---|---|
| CRP | 0.64 (0.58-0.70) | 0.59 (0.49-0.69) | 0.48 (0.36-0.59) | 0.70 (0.58-0.81) | 0.83 (0.73-0.92) | 0.58 (0.52-0.65) |
| IP-10* | 0.64 (0.58-0.69) | 0.54 (0.43-0.65) | 0.69 (0.61-0.78) | 0.64 (0.55-0.74) | 0.65 (0.51-0.78) | 0.63 (0.57-0.69) |
| IL-10 | 0.63 (0.58-0.69) | 0.76 (0.67-0.85) | 0.58 (0.49-0.68) | 0.61 (0.49-0.72) | 0.60 (0.49-0.73) | 0.64 (0.58-0.70) |
| Hb* | 0.60 (0.54-0.67) | 0.65 (0.50-0.80) | 0.59 (0.49-0.70) | 0.67 (0.55-0.78) | 0.46 (0.33-0.60) | 0.64 (0.57-0.71) |
| Ang-1* | 0.48 (0.42-0.53) | 0.48 (0.38-0.59) | 0.63 (0.61-0.78) | 0.43 (0.32-0.54) | 0.37 (0.26-0.48) | 0.51 (0.45-0.57) |

*Reciprocal concentration used, as lower biomarker concentrations known to be indicative of more severe disease.

Missing data: Ang-1, Ang-2, sFlt-1, IL-6, IL-8, IL-10, IP-10, PCT, sTNFR-1, sTREM-1, n = 91; CHI3L1, n = 93; CRP, n = 96; IL-1ra, n = 92; suPAR, n = 124; lactate, glucose, n = 98; Hb, n = 574; WHO Danger Sign, n = 7, LqSOFA, n = 3, SIRS, n = 578.

### 16. Table S14a: Participants who received parenteral treatment prior to baseline data collection.

| | Clinical parameters | Blood collection |
|---|---|---|
| **Proportion** [1, a] | 301 / 4,405 (8.8%) | 340 / 3,405 (10.0%) |
| **Time (minutes)** [2, b] | 6.0 (4.0, 10.0) | 30.0 (15.0, 50.0) |

[1]Median (IQR); [2] n / N (%)

[a] proportion of participants with data collection after initiation of parenteral treatment; [b] time between parenteral treatment initiation and data collection.

**Table S14b: Prognostic performance of endothelial and immune activation markers and clinical assessment tools to predict progression to severe disease within two days of enrolment in participants that did not receive parenteral treatment prior to baseline data collection.**

| | **AUC (95% CI)**<br>N = events / non-events | |
|---|---|---|
| | **Overall**<br>N = 133 / 3,272 | **Sensitivity analysis**<br>N = 55 / 2,982 |
| **Clinical assessment tools** | | |
| WHO Danger Sign | 0.75 (0.71-0.80) | 0.76 (0.69-0.82) |
| LqSOFA | 0.74 (0.69-0.78) | 0.67 (0.60-0.74) |
| SIRS | 0.63 (0.58-0.68) | 0.66 (0.58-0.74) |
| **Biomarkers** | | |
| sTREM-1 | 0.86 (0.82-0.90) | 0.82 (0.76-0.88) |
| sFlt-1 | 0.81 (0.77-0.86) | 0.72 (0.64-0.80) |
| Ang-2 | 0.80 (0.75-0.85) | 0.68 (0.59-0.77) |
| PCT | 0.80 (0.75-0.84) | 0.77 (0.70-0.84) |
| sTNFR-1 | 0.77 (0.72-0.82) | 0.76 (0.68-0.84) |
| IL-8 | 0.76 (0.70-0.81) | 0.70 (0.61-0.79) |
| IL-6 | 0.74 (0.69-0.79) | 0.82 (0.76-0.88) |
| CHI3L1 | 0.74 (0.68-0.79) | 0.73 (0.65-0.81) |
| suPAR | 0.69 (0.64-0.75) | 0.60 (0.50-0.69) |
| Lactate | 0.68 (0.62-0.74) | 0.55 (0.46-0.64) |
| Glucose | 0.67 (0.61-0.73) | 0.62 (0.53-0.71) |
| IL-1ra | 0.66 (0.60-0.72) | 0.70 (0.63-0.77) |
| CRP | 0.64 (0.58-0.70) | 0.73 (0.65-0.80) |
| IP-10* | 0.64 (0.58-0.69) | 0.58 (0.48-0.66) |
| IL-10 | 0.63 (0.58-0.69) | 0.65 (0.57-0.73) |
| Hb* | 0.60 (0.54-0.67) | 0.49 (0.39-0.58) |
| Ang-1* | 0.48 (0.42-0.53) | 0.50 (0.42-0.58) |

*Reciprocal concentration used, as lower biomarker concentrations known to be indicative of more severe disease.

Missing data: Ang-1, Ang-2, sFlt-1, IL-6, IL-8, IL-10, IP-10, PCT, sTNFR-1, sTREM-1, n = 91; CHI3L1, n = 93; CRP, n = 96; IL-1ra, n = 92; suPAR, n = 124; lactate, glucose, n = 98; Hb, n = 574; WHO Danger Sign, n = 7, LqSOFA, n = 3, SIRS, n = 578.

### 17. Table S15: STROBE checklist.

| | Item No | Recommendation | Page No |
|---|---|---|---|
| **Title and abstract** | 1 | (*a*) Indicate the study's design with a commonly used term in the title or the abstract<br>(*b*) Provide in the abstract an informative and balanced summary of what was done and what was found | 1<br>3 |
| **Introduction** | | | |
| Background/rationale | 2 | Explain the scientific background and rationale for the investigation being reported | 7 |
| Objectives | 3 | State specific objectives, including any prespecified hypotheses | 7 |
| **Methods** | | | |
| Study design | 4 | Present key elements of study design early in the paper | 8 |
| Setting | 5 | Describe the setting, locations, and relevant dates, including periods of recruitment, exposure, follow-up, and data collection | 8, appendix 2-3 |
| Participants | 6 | (*a*) Give the eligibility criteria, and the sources and methods of selection of participants. Describe methods of follow-up<br>(*b*) For matched studies, give matching criteria and number of exposed and unexposed | 8-9<br>NA |
| Variables | 7 | Clearly define all outcomes, exposures, predictors, potential confounders, and effect modifiers. Give diagnostic criteria, if applicable | 9-11 |
| Data sources/ measurement | 8* | For each variable of interest, give sources of data and details of methods of assessment (measurement). Describe comparability of assessment methods if there is more than one group | 9-11, appendix 9 |
| Bias | 9 | Describe any efforts to address potential sources of bias | 8, appendix 4-5 |
| Study size | 10 | Explain how the study size was arrived at | 12 |
| Quantitative variables | 11 | Explain how quantitative variables were handled in the analyses. If applicable, describe which groupings were chosen and why | 12 |
| Statistical methods | 12 | (*a*) Describe all statistical methods, including those used to control for confounding<br>(*b*) Describe any methods used to examine subgroups and interactions<br>(*c*) Explain how missing data were addressed<br>(*d*) If applicable, explain how loss to follow-up was addressed<br>(*e*) Describe any sensitivity analyses | 12, 16 |
| **Results** | | | |
| Participants | 13* | (a) Report numbers of individuals at each stage of study—eg numbers potentially eligible, examined for eligibility, confirmed eligible, included in the study, completing follow-up, and analysed<br>(b) Give reasons for non-participation at each stage<br>(c) Consider use of a flow diagram | Appendix 11 |
| Descriptive data | 14* | (a) Give characteristics of study participants (eg demographic, clinical, social) and information on exposures and potential confounders<br>(b) Indicate number of participants with missing data for each variable of interest<br>(c) Summarise follow-up time (eg, average and total amount) | 13<br>Table 1<br>Table S8<br>Table S9<br>Table S10 |
| Outcome data | 15* | Report numbers of outcome events or summary measures over time | 13 |

| | | | |
|---|---|---|---|
| Main results | 16 | (*a*) Give unadjusted estimates and, if applicable, confounder-adjusted estimates and their precision (eg, 95% confidence interval). Make clear which confounders were adjusted for and why they were included<br>(*b*) Report category boundaries when continuous variables were categorized<br>(*c*) If relevant, consider translating estimates of relative risk into absolute risk for a meaningful time period | 14 |
| Other analyses | 17 | Report other analyses done—eg analyses of subgroups and interactions, and sensitivity analyses | 14-16 |
| **Discussion** | | | |
| Key results | 18 | Summarise key results with reference to study objectives | 17 |
| Limitations | 19 | Discuss limitations of the study, taking into account sources of potential bias or imprecision. Discuss both direction and magnitude of any potential bias | 18-19 |
| Interpretation | 20 | Give a cautious overall interpretation of results considering objectives, limitations, multiplicity of analyses, results from similar studies, and other relevant evidence | 19-20 |
| Generalisability | 21 | Discuss the generalisability (external validity) of the study results | 18-19 |
| **Other information** | | | |
| Funding | 22 | Give the source of funding and the role of the funders for the present study and, if applicable, for the original study on which the present article is based | 4,12 |

### 18. Table S16: REMARK checklist.

| | Item to be reported | Page no. |
|---|---|---|
| **INTRODUCTION** | | |
| 1 | State the marker examined, the study objectives, and any pre-specified hypotheses. | 5, 9-10 |
| **MATERIALS AND METHODS** | | |
| *Patients* | | |
| 2 | Describe the characteristics (e.g., disease stage or co-morbidities) of the study patients, including their source and inclusion and exclusion criteria. | 8 |
| 3 | Describe treatments received and how chosen (e.g., randomized or rule-based). | 9 |
| *Specimen characteristics* | | |
| 4 | Describe type of biological material used (including control samples) and methods of preservation and storage. | 10 |
| *Assay methods* | | |
| 5 | Specify the assay method used and provide (or reference) a detailed protocol, including specific reagents or kits used, quality control procedures, reproducibility assessments, quantitation methods, and scoring and reporting protocols. Specify whether and how assays were performed blinded to the study endpoint. | 10-11, appendix 10 |
| *Study design* | | |
| 6 | State the method of case selection, including whether prospective or retrospective and whether stratification or matching (e.g., by stage of disease or age) was used. Specify the time period from which cases were taken, the end of the follow-up period, and the median follow-up time. | 8-9, appendix 2-5 |
| 7 | Precisely define all clinical endpoints examined. | 11 |
| 8 | List all candidate variables initially examined or considered for inclusion in models. | 9-10 |
| 9 | Give rationale for sample size; if the study was designed to detect a specified effect size, give the target power and effect size. | 12 |
| *Statistical analysis methods* | | |
| 10 | Specify all statistical methods, including details of any variable selection procedures and other model-building issues, how model assumptions were verified, and how missing data were handled. | 12 |
| 11 | Clarify how marker values were handled in the analyses; if relevant, describe methods used for cutpoint determination. | Table 2 |
| **RESULTS** | | |
| *Data* | | |
| 12 | Describe the flow of patients through the study, including the number of patients included in each stage of the analysis (a diagram may be helpful) and reasons for dropout. Specifically, both overall and for each subgroup extensively examined report the numbers of patients and the number of events. | Appendix 11 |
| 13 | Report distributions of basic demographic characteristics (at least age and sex), standard (disease-specific) prognostic variables, and tumor marker, including numbers of missing values. | 12, Table 1, Table S8-10 |
| *Analysis and presentation* | | |
| 14 | Show the relation of the marker to standard prognostic variables. | 14, Figure 2 |
| 15 | Present univariable analyses showing the relation between the marker and outcome, with the estimated effect (e.g., hazard ratio and survival probability). Preferably provide similar analyses for all other variables being analyzed. For the effect of a tumor marker on a time-to-event outcome, a Kaplan-Meier plot is recommended. | Figure 2 |
| 16 | For key multivariable analyses, report estimated effects (e.g., hazard ratio) with confidence intervals for the marker and, at least for the final model, all other variables in the model. | NA |
| 17 | Among reported results, provide estimated effects with confidence intervals from an analysis in which the marker and standard prognostic variables are included, regardless of their statistical significance. | 14, Figure 2 |
| 18 | If done, report results of further investigations, such as checking assumptions, sensitivity analyses, and internal validation. | 14-16 |
| **DISCUSSION** | | |
| 19 | Interpret the results in the context of the pre-specified hypotheses and other relevant studies; include a discussion of limitations of the study. | 17-19 |
| 20 | Discuss implications for future research and clinical value. | 19-20 |

### 19. Table S17. Spot Sepsis Investigator Group.

| Name | Affiliation |
|---|---|
| **Mohammad Yazid Abdad** | Mahidol-Oxford Tropical Medicine Research Unit, Mahidol University, Thailand; Centre for Tropical Medicine and Global Health, University of Oxford, UK |
| **Riris Andono Ahmad** | Centre for Tropical Medicine, Universitas Gadjah Mada, Indonesia |
| **Dinh Thi Van Anh** | Viet Nam National Children's Hospital, Viet Nam |
| **Eggi Arguni** | Centre for Tropical Medicine, Universitas Gadjah Mada, Indonesia |
| **Elizabeth A Ashley** | Lao-Oxford-Mahosot Hospital-Wellcome Trust Research Unit, Mahosot Hospital, Laos; Centre for Tropical Medicine and Global Health, University of Oxford, UK |
| **Elizabeth M Batty** | Mahidol-Oxford Tropical Medicine Research Unit, Mahidol University, Thailand; Centre for Tropical Medicine and Global Health, University of Oxford, UK |
| **Latsaniphone Boutthasavong** | Lao-Oxford-Mahosot Hospital-Wellcome Trust Research Unit, Mahosot Hospital, Laos |
| **Sakib Burza** | Médecins Sans Frontières Operational Centre Barcelona, Spain; Clinical Research Department, London School of Hygiene and Tropical Medicine, UK; Health in Harmony, United States of America |
| **Arjun Chandna** | Cambodia Oxford Medical Research Unit, Angkor Hospital for Children, Cambodia; Centre for Tropical Medicine and Global Health, University of Oxford, UK |
| **Ngoun Chanpheaktra** | Angkor Hospital for Children, Cambodia |
| **Tran Quoc Dat** | Vietnam National Children's Hospital, Viet Nam |
| **Vu Quoc Dat** | Hanoi Medical University, Viet Nam |
| **Nicholas P J Day** | Mahidol-Oxford Tropical Medicine Research Unit, Mahidol University, Thailand; Centre for Tropical Medicine and Global Health, University of Oxford, UK |
| **Arjen M Dondorp** | Mahidol-Oxford Tropical Medicine Research Unit, Mahidol University, Thailand; Centre for Tropical Medicine and Global Health, University of Oxford, UK |
| **Prakash Ghosh** | International Centre for Diarrhoeal Disease Research, Bangladesh; Institute of Animal Hygiene and Veterinary Public Health, Leipzig University, Germany; Technische Universität Berlin, Germany |
| **Carolina Jimenez** | Médecins Sans Frontières Operational Centre Barcelona, Spain |
| **Kevin Kain** | Department of Laboratory Medicine and Pathobiology, University of Toronto, Canada |
| **Muhammad Karyana** | INA-RESPOND, Ministry of Health Republic Indonesia, Indonesia |
| **Suy Keang** | Angkor Hospital for Children, Cambodia; Cambodia Oxford Medical Research Unit, Angkor Hospital for Children, Cambodia |
| **Sommay Keomany** | Salavan Provincial Hospital, Laos |
| **Rungnapa Khamboocha** | Mahidol-Oxford Tropical Medicine Research Unit, Mahidol University, Thailand |
| **Constantinos Koshiaris** | Department of Primary Care Health Sciences, University of Oxford, UK |
| **Khamfong Kunlaya** | Lao-Oxford-Mahosot Hospital-Wellcome Trust Research Unit, Mahosot Hospital, Laos |
| **Estrella Lasry** | Médecins Sans Frontières Operational Centre Barcelona, Spain |

| | |
|---|---|
| **Bui Thanh Liem** | University of Medicine and Pharmacy at Ho Chi Minh City, Vietnam |
| **Nguyen Huy Luan** | University of Medicine and Pharmacy at Ho Chi Minh City, Vietnam |
| **Yoel Lubell** | Mahidol-Oxford Tropical Medicine Research Unit, Mahidol University, Thailand; Centre for Tropical Medicine and Global Health, University of Oxford, UK; Amsterdam Institute of Global Health and Development, The Netherlands |
| **Raman Mahajan** | Médecins Sans Frontières Operational Centre Barcelona, Spain |
| **Saysamone Malavong** | Savannakhet Provincial Hospital, Laos |
| **Mayfong Mayxay** | Lao-Oxford-Mahosot Hospital-Wellcome Trust Research Unit, Mahosot Hospital, Laos; Institute for Research and Education Development, University of Health Sciences, Laos; Centre for Tropical Medicine and Global Health, University of Oxford, UK |
| **Chonticha Menggred** | Mahidol-Oxford Tropical Medicine Research Unit, Mahidol University, Thailand |
| **Dinesh Mondal** | International Centre for Diarrhoeal Disease Research, Bangladesh |
| **Phung Nguyen The Nguyen** | University of Medicine and Pharmacy at Ho Chi Minh City, Vietnam |
| **Rafael Perera-Salazar** | Department of Primary Care Health Sciences, University of Oxford, UK |
| **Chom Phaiphichit** | Lao-Oxford-Mahosot Hospital-Wellcome Trust Research Unit, Mahosot Hospital, Laos |
| **Chanthala Phamisith** | Savannakhet Provincial Hospital, Laos |
| **Phan Huu Phuc** | Viet Nam National Children's Hospital, Viet Nam |
| **Tiengkham Pongvongsa** | Savannakhet Provincial Health Office, Laos |
| **Sayaphet Rattanavong** | Lao-Oxford-Mahosot Hospital-Wellcome Trust Research Unit, Mahosot Hospital, Laos |
| **Michael Rekart** | Médecins Sans Frontières Operational Centre Barcelona, Spain |
| **Melissa Richard-Greenblatt** | Centre for Tropical Medicine and Global Health, University of Oxford, UK; Department of Laboratory Medicine and Pathobiology, University of Toronto, Canada; Hospital for Sick Children, Canada |
| **Bran Sambou** | Cambodia Oxford Medical Research Unit, Angkor Hospital for Children, Cambodia |
| **Khalid Shams** | Médecins Sans Frontières Operational Centre Barcelona, Spain |
| **Mohammad Shomik** | International Centre for Diarrhoeal Disease Research, Bangladesh |
| **Phouthalavanh Souvannasing** | Salavan Provincial Hospital, Laos |
| **Phattaranit Tanunchai** | Mahidol-Oxford Tropical Medicine Research Unit, Mahidol University, Thailand |
| **Janjira Thaipadungpanit** | Mahidol-Oxford Tropical Medicine Research Unit, Mahidol University, Thailand; Faculty of Tropical Medicine, Mahidol University, Thailand |
| **Watcharintorn Thongpiam** | Mahidol-Oxford Tropical Medicine Research Unit, Mahidol University, Thailand |
| **Claudia Turner** | Angkor Hospital for Children, Cambodia; Cambodia Oxford Medical Research Unit, Angkor Hospital for Children, Cambodia; Centre for Tropical Medicine and Global Health, University of Oxford, UK |
| **Paul Turner** | Cambodia Oxford Medical Research Unit, Angkor Hospital for Children, Cambodia; Centre for Tropical Medicine and Global Health, University of Oxford, UK |

| | |
|---|---|
| **Souphaphone Vannachone** | Lao-Oxford-Mahosot Hospital-Wellcome Trust Research Unit, Mahosot Hospital, Laos |
| **Asama Vinitsorn** | Mahidol-Oxford Tropical Medicine Research Unit, Mahidol University, Thailand |
| **Ranitha Vongpromek** | Mahidol-Oxford Tropical Medicine Research Unit, Mahidol University, Thailand |
| **Manivanh Vongsouvath** | Lao-Oxford-Mahosot Hospital-Wellcome Trust Research Unit, Mahosot Hospital, Laos |
| **Naomi Waithira** | Mahidol-Oxford Tropical Medicine Research Unit, Mahidol University, Thailand; Centre for Tropical Medicine and Global Health, University of Oxford, UK |
| **James A Watson** | Mahidol-Oxford Tropical Medicine Research Unit, Mahidol University, Thailand; Centre for Tropical Medicine and Global Health, University of Oxford, UK |
| **Mikhael Yosia** | Médecins Sans Frontières Operational Centre Barcelona, Spain |
| **Asri Yuniastuti** | Wates District Hospital, Indonesia |
